# Tracing and testing multiple generations of contacts to COVID-19 cases: cost-benefit tradeoffs

**DOI:** 10.1101/2021.06.29.21259723

**Authors:** Jungyeol Kim, Xingran Chen, Hesam Nikpey, Harvey Rubin, Shirin Saeedi Bidokhti, Saswati Sarkar

## Abstract

Traditional contact tracing for COVID-19 tests the direct contacts of those who test positive even if the contacts do not show any symptom. But, by the time an infected individual is tested, the infection starting from the person may have infected a chain of individuals. Hence, why should the testing stop at direct contacts, and not test secondary, tertiary contacts or even contacts further down? One deterrent in testing long chains of individuals right away may be that it substantially increases the testing load, or does it? We investigate the costs and benefits of such multi-hop contact tracing for different number of hops. Considering a large number of contact topologies, spanning synthetic networks of divergent characteristics and those constructed from recorded interactions, we show that the cost-benefit tradeoff can be characterized in terms of a single measurable attribute, the *initial epidemic growth rate*. Once this growth rate crosses a threshold, multi-hop contact tracing substantially reduces the outbreak size compared to traditional contact tracing. Multi-hop even incurs a lower cost compared to the traditional contact tracing for a large range of values of the growth rate. The cost-benefit tradeoffs and the choice of the number of hops can be classified into three phases, with sharp transitions between them, depending on the value of the growth rate. The need for choosing a larger number of hops becomes greater as the growth rate increases or the environment becomes less conducive toward containing the disease.

**Author summary:** The COVID-19 pandemic has wrecked havoc on lives and livelihoods worldwide. Other epidemics may well emerge in future and one needs a preparedness to prevent their growth into another pandemic. During the early stages of a new epidemic, or even a mutated version of an earlier epidemic, pharmaceutical interventions may not be available but contact tracing and timely quarantine are among the few available control measures. We show that 1) traditional contact tracing may not successfully contain the outbreak depending on the rate of growth of the epidemic, but 2) the cost-benefit tradeoffs may be substantially enhanced through the deployment of a natural multi-hop generalization which tests contact chains starting from those who test positive.

## Introduction

To slow down the spread of COVID-19, public health authorities like the US Center for Disease Control and Prevention (CDC) recommended testing those who have in the recent past been in physical proximity with an individual who has tested positive, even when the contacts do not exhibit any symptom [1]. This preemptive action, commonly known as contact tracing, is deployed because given how contagious the disease is, a patient is likely to have passed the virus to their contacts, and the infected contacts have the potential to infect others even before they show symptoms [2]. Discovering and quarantining those infected contacts will stop them from spreading the disease much earlier than a strategy in which only symptomatic individuals who seek medical help are tested. Slowing down the spread by contact tracing comes at the cost of an increase in the testing load, yet, the cost-benefit tradeoff for contact tracing is understood to be substantially favorable, as compared to universal lockdowns for example, which has led to economic downturns in several countries.

In this paper, we want to understand under what circumstances traditional contact tracing alone is sufficient to contain the virus and why such containment is attainable in those circumstances. We also want to understand circumstances where the traditional approach is not efficient enough and how we can overcome this. A question that naturally arises in this regard is if cost-benefit tradeoffs may be enhanced through natural generalizations of the core concept of contact tracing - this is what we seek to answer in this paper. In the time that elapses between when an individual, I, is infected until I is tested, the disease spreads from I through a chain of several hops - I infects those I is in contact with, those whom I infects can infect their contacts, the infected contacts can infect their contacts, and so on. A recent study suggests that, due to the high speed of transmission, the epidemic may continue to grow even if all contacts are quarantined with some delay [3].

Fewer people are likely to be infected by testing and quarantining not only direct contacts of an individual who tests positive, but contacts of the contacts and so on (Fig 1a). Such *multi-hop* tracing and testing will enable identification and quarantine of the individuals further down the chain who have been exposed, earlier than if we had tested only the direct contacts of those who have tested positive and then reach down the chain iteratively. To see why multi-hop contact tracing may be effective note that an infectious disease spreads through growth of clusters of infected individuals around one or more origins, e.g., during the spread of COVID-19 large clusters were observed in meat-packing plants in seven countries, and an e-commerce distribution warehouse in South Korea [4]. Contact tracing also form clusters of tested individuals that grow from and around one or more individuals who initially test positive (Fig 1b). In this sense, contact tracing emulates the spread of the disease. If the testing cluster grows faster than the infection cluster and also substantially overlaps with the latter, the outbreak will be contained. And by virtue of its design, multi-hop contact tracing grows the testing cluster faster than traditional contact tracing.

**Fig 1.**
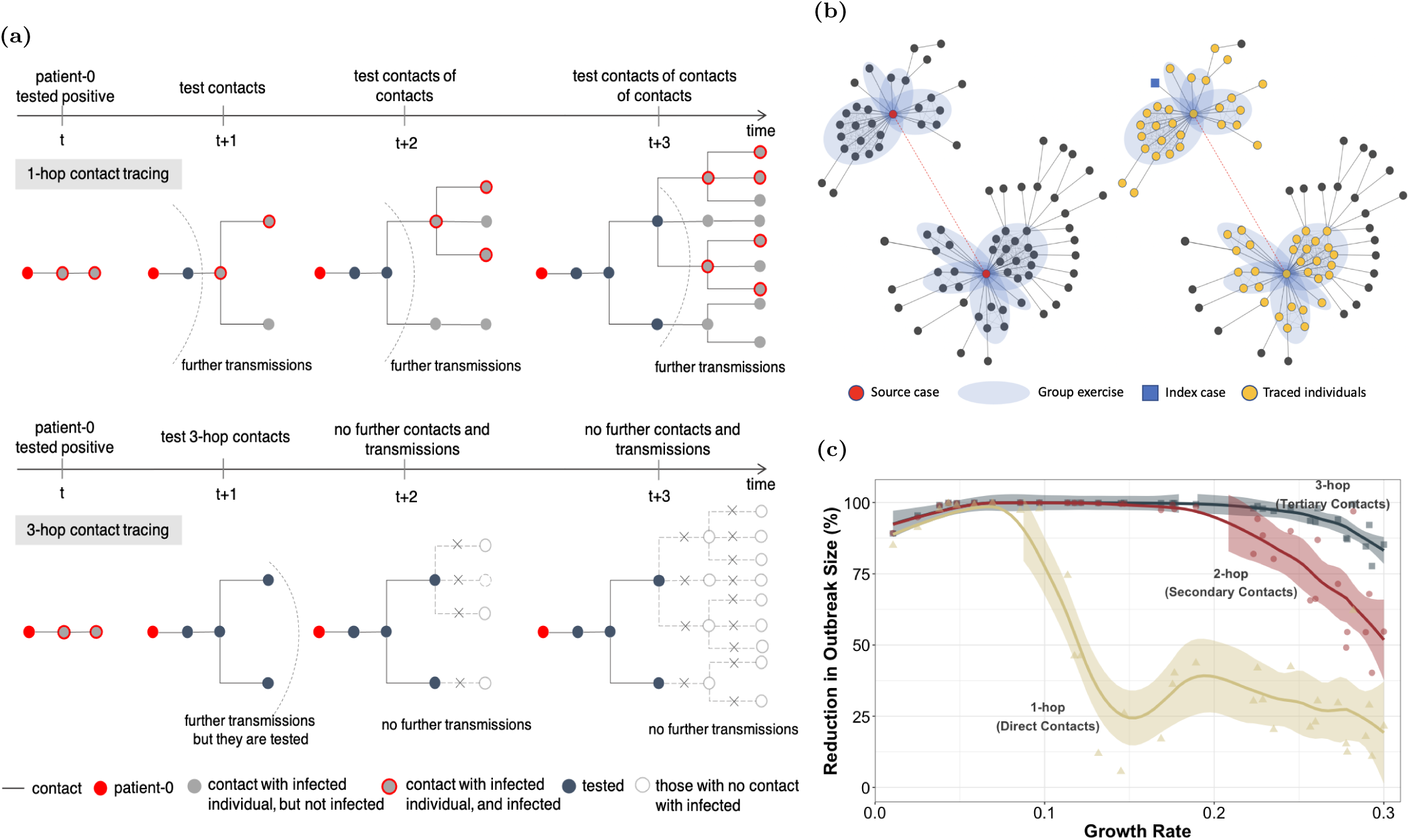
Multi-hop contact tracing illustration and the effectiveness of the tracing scheme. (a) We use an example to illustrate and compare 1-hop contact tracing shown on top (i.e., tracing and testing only the direct contacts of those who test positive) and 3-hop contact tracing shown on bottom (i.e., tracing and testing the direct, secondary and tertiary contacts of those who test positive). Here, at time *t* when patient-0 (red) is tested by a health authority, the infection has already propagated 2 hops. By time *t* + 3, both tracing policies test 4 individuals (marked in black) other than the patient-0; the 3-hop policy tests and quarantines the positive ones in a shorter time, while 1-hop tests and quarantines them progressively and therefore over longer times. Accordingly, only 3 individuals are infected under the 3-hop policy, while 10 individuals are infected under the 1-hop policy. (b) Left network: This figure is a partial network based on epidemiological investigation information by the Korea Centers for Disease Control and Prevention (KCDC) and local governments [5]. It illustrates how infection spreads from two dance instructors (source cases; red circle), both of whom attended a workshop on Feb 15, 2020, in South Korea. Subsequently they separately taught dance classes indoor and spread to the attendees who spread to their contacts. The blue shaded area represents the instructors and the attendees in each dance class, and the close contacts (gray dashed line edges) among them in the class. The gray solid lines represent the contact during which the disease is transmitted. The dashed red line between the two instructors indicate that they were in contact (because they simultaneously attended the workshop). Right network: Suppose an index case indicated by the blue square is identified by the health authority. When multi-hop (e.g., 3 hop) contact tracing is done, the traced nodes also form a cluster. Thus, intuitively, the growth of the traced cluster emulates the growth of the infected cluster, this emulation helps in containment. (c) The percentage of reduction in the number of infections when 1-hop, 2-hop, and 3-hop contact tracing policies were performed, compared to when no contact tracing was performed, as a function of the growth rate. For example, a point where the y-axis corresponds to 20% indicates that the number of infections that would occur when no contact tracing is performed can be reduced by 20% through the implementation of the contact tracing policy. Tracing secondary (2-hop) and tertiary (3-hop) contacts can reduce the outbreak size significantly in cases of high growth rate where tracing direct (1-hop) contacts alone cannot sufficiently reduce the outbreak size. The growth rate (x-axis) characterizes the intrinsic speed of virus spread in the absence of any public health intervention, which is determined by contact patterns and transmission probabilities. Data points having the same value of the growth rate represent that the virus spreads under the same contact pattern and the same transmission probability. The solid lines correspond to the LOESS smoother with a span value of 0.3 and the shadings represent the 95% confidence interval around the smoother line. Refer to Model Dynamics section for details on the contact networks and transmission probabilities we use.

For these reasons, depending on the rate of growth of the epidemic, traditional contact tracing may not successfully contain the outbreak. Note that, as shown in Fig 1c, our simulations in a large number of large-scale contact networks reveal that tracing direct contacts alone cannot sufficiently contain the virus when the rate of growth of the epidemic is beyond a threshold, even under the ideal condition of full compliance with contact tracing and quarantine. On the other hand, tracing secondary and tertiary contacts can reduce the outbreak size significantly despite the high rate of growth of the epidemic.

Multi-hop tracing strategies have been sporadically deployed in practice with considerable success. For example, in Vietnam, public health authority sometimes reached out to tertiary contacts, and found and tested as many as 200 contacts for each case [6]; many of those who were traced and quarantined during the first 100 days of the pandemic were in fact secondary contacts of those who tested positive [7]. Vietnam reported only a total of 1, 465 PCR-confirmed cases and 35 deaths by the end of 2020 [8]. Nonetheless, this concept has not been comprehensively and systemically investigated - this is the void this paper seeks to fill in.

The following questions arise in context of multi-hop tracing: 1) Under what circumstances can traditional contact tracing not significantly reduce the outbreak size? In these cases, do aggressive preemptive tracing schemes under multi-hop reduce the outbreak size significantly? 2) Do such schemes necessarily increase the overall number of tests and quarantines? The answer is not a priori clear as reduction in overall infection spread through such a strategy may eventually reduce the number of tests required, as illustrated in Fig 1a. 3) If multi-hop tracing turns out to be beneficial, how many hops provide the best cost-benefit tradeoff? Does a *saturation phenomenon* in which the benefit increases only marginally by increasing the number of hops beyond a certain point arise? If so, what is the saturation point? 4) How do these answers depend on the attributes of the tests, the time at which contact tracing is deployed, and the behavioral dynamics, that is, the extent of public compliance of public health directives? We proceed to answer these questions in this paper.

We formalize the aggressive preemptive tracing and testing scheme as *k-hop contact tracing*, where *k* is the depth of the contact chain that is traced. For example, *k* = 0 does not trace contacts and tests only those who show symptoms and seek medical help, *k* = 1 is the traditional contact tracing that tests the direct contacts of an individual who tests positive, *k* = 2 additionally tests the contacts of the contacts, *k* = 3 tests yet another hop of contacts, and so on. We call the multiple generations of contacts to COVID-19 cases (i.e., *k*-hop contact tracing for *k* ≥ 2) *multi-hop contact tracing*.

We quantify the costs and benefit of contact tracing over a course of 6 months (180 days) starting from the day after contact tracing is initiated, and compare the results for multi-hop contact tracing with 1-hop contact tracing. The *benefit* is defined as the percentage of reduction in the number of infections over the period compared to when no contact tracing was performed. The *costs* are comprised of 1) the number of tests and 2) total sum of days of quarantine for the entire population over the period.

However, the nature of the cost-benefit tradeoff for multi-hop contact tracing depends on practical aspects of contact tracing and testing that are present in all types of infectious diseases. First of all, behavioral dynamics undermine the efficacy of contact tracing. Individuals do not always cooperate with public-health authorities by 1) disclosing their contacts 2) quarantining when exposed to those who test positive. Secondly, the tests suffer from false negatives and false positives. If an individual tests negative falsely, his *k*-hop contacts will not be traced and tested (unless those contacts are within *k*-hop of another individual who tests positive). This undermines the ability of the tracing strategy to contain the outbreak. False positives may increase cost by setting off a chain of unnecessary tracing and testing. Thirdly, tests can have different turnaround times, high turnaround times delay tracing the contacts of those infected. Lastly, contact tracing in its entirety may be initiated by public health officials only after the infection level in the target populace crosses a certain threshold. All these attributes are likely to affect the outcome of the tracing. These attributes depend on regional and cultural characteristics and public health policies which are different in different ambiences. Given the inherent uncertainty of the settings and the heterogeneity for different venues, we consider a range of values of the above attributes based on estimates available in the literature.

The dynamics of epidemic spread are governed by inter-personal contact patterns and probability with which a contagious individual infects a susceptible individual in an interaction. Thus, these factors uniquely determine *initial epidemic growth rate* that characterizes the intrinsic speed of virus spread within each community in the absence of any public health intervention. We consider diverse large-scale contact networks and a range of values of the transmission probability.

Under a variety of contact patterns and transmission probabilities, our simulations reveal that the nature of the cost-benefit tradeoff for multi-hop contact tracing can be characterized in terms of the growth rate, and the nature remains largely stable to variation of the above-mentioned practical aspects in reasonable ranges. When the growth rate is low, 1-hop contact tracing alone can sufficiently contain the virus. However, once the growth rate crosses a threshold value, a sharp *phase transition* is exhibited. Specifically, at intermediate growth rates, the benefit that 1-hop provides dramatically decreases as compared to the low growth rate range, and multi-hop contact tracing offers substantial further benefit *even at a lower cost compared to* 1*-hop*. At high growth rates, multi-hop contact tracing provides substantial further benefit but incurs greater costs, as compared to 1-hop contact tracing.

Our results also reveal that the further benefit of adding more hops beyond 1-hop tends to diminish progressively in most settings, thus a *saturation phenomenon* is observed throughout. In general, the hop number at which the saturation phenomenon is observed becomes greater as the growth rate increases and/or the environments regarding practical aspects becomes less conducive toward containing the disease. Specifically, increasing the number of hops beyond 3-hop provides only marginal benefit in most cases, thus the saturation point is confined to 1, 2, and 3 hops, despite variations of all the above-mentioned attributes; the need for considering 4 and 5 hops largely arises for very limited conditions such as higher growth rates and more challenging environments.

Note that, in this study, we consider COVID-19 as an example of an infectious epidemic. Infectious diseases primarily vary in the stages of the disease evolution, and in the parameters governing their durations and transmission probability. In general, contact tracing (and quarantine) may be used to contain every infectious epidemic, and, in fact, constitutes one of the few available mechanisms to contain the outbreak during the early phases of a new epidemic when pharmaceutical interventions are often not available. This can prevent an epidemic from developing into a pandemic with an enormous toll like that COVID-19 imposed. Wherever contact tracing may be deployed, multi-hop contact tracing can be deployed too. And, our framework for assessing the cost and benefit tradeoffs of multi-hop contact tracing may be generalized to other infectious diseases by appropriately choosing the parameters for the disease and stages of the disease evolution. Our framework lends itself to such a generalization also because we have taken into account the attributes involving contact patterns, transmission probability, and practical aspects of contact tracing and testing that can apply to all kinds of infectious diseases.

Before presenting our main results, we discuss some of the challenges that the real-world implementation of multi-hop contract tracing may face. Undoubtedly, manual tracing of contacts is a labor intensive task even for single-hop tracing. It requires large numbers of trained public health workers to call people who have tested positive and their contacts, and to provide information as necessary. This over-load may thus be exacerbated for multi-hop contact tracing. In terms of the total over-load (averaged over time), we determine regimes of operation (in terms of the growth rate of the disease process) for which the over-load is comparable/less/larger than one-hop contact tracing. In terms of the daily over-load, we expect a larger over-load especially during the initial days of the tracing. This is because although in many instances testing and quarantining (and therefore tracing) costs over time were lower for multi-hop as compared to single-hop, at the start, invariably multi-hop needs to trace more than single-hop. These challenges can, however, be overcome by digital contact tracing apps, albeit various challenges and concerns need to be addressed. For example, in democracies the digitization is often critically reliant on the willingness of the populace to download the apps the health authorities use, which has again varied from country to country for the COVID-19 outbreak. For example, in Singapore over 92% of the population over 6 years of age had downloaded the governmental contact tracing app on their smartphone [9], but the fraction has been lower in many other countries, particularly those in Europe and US. In this study, we assume the implementation of digital contact tracing by the target populace, but also consider scenarios in which cooperation on digital contact tracing is less universal.

## Model Dynamics and Contact Tracing Process

We consider a discrete time stochastic evolution of COVID-19 in a population that initially consists of susceptible and a few contagious individuals. We model the progression of the disease using a compartmental model (Fig 2b). The disease spreads from the contagious individual (CI) to the susceptible individual (SI) through mutual interaction. In any given interaction with a CI, an SI is infected with a probability *β*. This transmission probability depends on a range of factors such as whether the individuals observe social distancing, wear protective equipment and varies from one venue to another. Considering [11, 12], we consider a wide range of values for *β*, namely *β*∈ [0.1, 0.3]. After a *latency period* (the presymptomatic-latent and asymptomatic-latent are in this latency period), the newly infected individuals become contagious. Specifically, at the end of the latency period, the individuals either become *presymptomatic* (the stage before exhibiting symptoms), or *asymptomatic* (that is, they never show symptoms). Presymptomatics proceed to become *symptomatics* in the next stage. After a random delay, symptomatics opt for seeking medical help and testing, and become *ready-to-test*. Presymptomatics, asymptomatics, symptomatics all however are contagious. Refer to Methods for details on the systems we consider and the parameters we choose.

**Fig 2.**
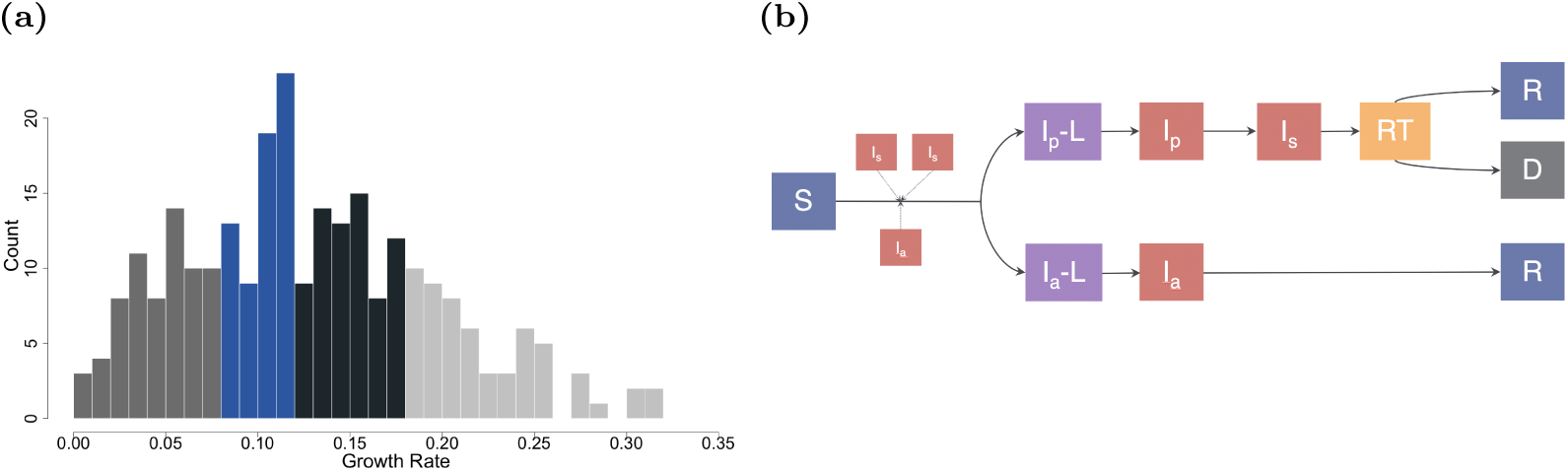
Real-world initial epidemic growth rates and virus transmission model illustration. (a) The real-world initial epidemic growth rates from 258 political units range from 0 to 0.31, with a median of 0.12 (interquartile range 0.08 - 0.17). Each quartile is filled with a different color. (b) Virus transmission model illustration. The Compartmental model consists of the following compartments: Susceptible (*S*), Presymptomatic-Latent (*I*_*p*_-*L*), Presymptomatic (*I*_*p*_), Symptomatic (*I*_*s*_), Ready-to-Test (*RT*), Asymptomatic-Latent (*I*_*a*_-*L*), Asymptomatic (*I*_*a*_), Recovered (*R*), and Dead (*D*).

Once the individual in question tests positive, the public health authority traces his *k*-hop contacts, over the last 14 days, and informs them at the end of the day that they may have been exposed under the assumption of implementation of digital contact tracing. Such contact tracing may be accomplished through digital contact tracing (refer to SI Appendix for details on digital contact tracing pertaining to multi-hop contact tracing). The authority asks them to self-quarantine for 14 days unless they are already under quarantine or ever tested positive before. We assume that the traced individuals are scheduled for testing in 3 days. The test results are available in 1 to 3 days. Individuals who test positive will not be tested again, but those who test negative can be tested again if they are traced again from an individual who tested positive.

We evaluate the multi-hop testing strategies through agent-driven simulation on diverse large-scale contact networks, spanning a large number of networks of a classical synthetic variety with *N* = 100, 000 individuals and empirical social contact networks. In the contact networks, the nodes represent the individuals and the edges their contacts; the degrees of the nodes represent the number of contacts of the corresponding individuals. Growth of an epidemic depends on structural attributes of the contact networks, such as 1) average path lengths between nodes 2) clustering coefficient 3) degree distribution. We consider two broad classes of synthetic networks, which captures different ranges of the above attributes: 1) Watts-Strogatz networks 2) Scale-free networks. By varying a parameter, referred to as the *rewiring probability*, of the Watts-Strogatz networks from 0 to 1, one can realize 1) average path lengths that range from linear to logarithmic functions of the number of nodes 2) clustering coefficients from high to vanishingly small [10]. Studies based on real data suggest that contact networks among individuals exhibit short (i.e., logarithmic) average path length and high clustering coefficients (commonly referred to as the small-world property) [13, 14]. When clustering coefficient is high, most of the contacts happen between individuals in given phases or clusters; when clustering coefficient is low, most contacts happen between randomly selected individuals. Both extremes and values in between can be captured by choosing the value of the rewiring probability [10]. The special case of the Watts-Strogatz model in which the average path length is logarithmic and the clustering coefficient is low corresponds to a variant of the Erdős Rényi random networks; we consider this variant as well. Scale-free networks exhibit heterogeneous degree distributions, i.e., the degree distribution has a high variance and only a polynomially decaying tail (‘fat-tailed’ distribution). The degree distribution in Watts-Strogatz models have exponentially decaying tails for usual choices of parameters. The implication of this difference is that scale-free networks invariably have some individuals with very high degree, while the probability of the same happening in Watts-Strogatz models is low.

Additionally, we consider a social contact network obtained from data recorded from social and professional interaction patterns that have been realized in practice. This data-set records interactions among 69, 441 individuals residing in 75 villages in the state of Karnataka in India [15]. In this data-set, each village consists of 354 - 1775 individuals in this data-set. The limitation of this data-set is that it contains information only on social interactions between individuals within each village. However, in reality, individuals living in different villages do come in contact, and pandemic spreads from one village to another through these contacts. Also, the cost-benefit trade-off for multi-hop contact tracing is best evaluated on large population sizes, otherwise the length of the contact chains will be limited by the size of the target populace. We therefore rewire *r*% of edges to introduce interactions between individuals of different villages, where *r* is a parameter.

By considering all of these diverse networks complementing each other in fundamental characteristics, we are able to assess the cost-benefit tradeoffs of multi-hop contact tracing and testing strategies for widely varying contact patterns. See Table 1 and Methods for details on all the synthetic and data-driven networks we consider.

**Table 1.** Contact Networks.

| Notation | Types of Networks | Population<br>$N$ | Average Degree<br>$\langle k \rangle$ | Diameter<br>$d$ | Average Path Length<br>$l$ | Average Clustering Coefficient<br>$\langle C \rangle$ |
| --- | --- | --- | --- | --- | --- | --- |
| WS1 | Watts-Strogatz network w/ $p = 0.01$ | 100,000 | 4.00 | 95 | 39.3 | 0.472 |
| WS2 | Watts-Strogatz network w/ $p = 0.1$ | 100,000 | 4.00 | 22 | 12.4 | 0.275 |
| WS3 | Watts-Strogatz network w/ $p = 1$ | 100,000 | 4.00 | 17 | 8.42 | 0.0000413 |
| WS4 | Watts-Strogatz network w/ $p = 0.01$ | 100,000 | 8.00 | 32 | 16.6 | 0.606 |
| WS5 | Watts-Strogatz network w/ $p = 0.1$ | 100,000 | 8.00 | 11 | 7.50 | 0.349 |
| WS6 | Watts-Strogatz network w/ $p = 1$ | 100,000 | 8.00 | 10 | 5.77 | 0.0000804 |
| SF | Scale-free network | 100,000 | 4.00 | 10 | 5.88 | 0.000651 |
| DATA1 | Data-driven network w/ $r = 1$ | 69,441 | 8.49 | 17 | 8.81 | 0.627 |
| DATA2 | Data-driven network w/ $r = 3$ | 69,441 | 8.49 | 15 | 7.45 | 0.589 |
| DATA3 | Data-driven network w/ $r = 5$ | 69,441 | 8.49 | 15 | 6.97 | 0.553 |

*Notes.* $p$ is the rewiring probability of Watts-Strogatz networks, and $r$ is the mixing parameter in data-driven network. The average degree is the average number of edges per node. The distance between a pair of nodes is the length of the shortest path between them. The diameter is the maximum value of this distance over all pairs of nodes. The average path length is the average of this distance over all pairs of nodes; only the lengths of the existing paths are considered and averaged. Clustering coefficient of a node $i$ , $C_i$ , is defined as the ratio between the actual number of links between the neighbors of $i$ and the maximum possible number of links between the neighbors of $i$ [10]. This is high if there exists a large number of edges in the neighborhood of $i$ . The average clustering coefficient, $\langle C \rangle$ , is the average of $C_i$ over all nodes $i$ . The average degree, average path length, and average clustering coefficient are rounded to three significant figures.

We consider an attribute called *initial epidemic growth rate*, or more simply the *growth rate*, that characterizes the intrinsic speed of virus spread in each network. This attribute depends on the network structure and the transmission probability *β*. Let *N*_*t*_ and *N*_0_ respectively be the cumulative number of infected individuals on day *t* and day *t*_0_ in a target region, where *t*_0_ represents the start and [*t*_0_, *t*] an initial period of the epidemic growth. Now, similar to [16], we define the growth rate in the target region as (ln *N*_*t*_ − ln *N*_0_)*/*(*t* − *t*_0_). We choose this expression (particularly the logarithmic functions) because the growth of infections during the initial period has been widely observed to be exponential for different epidemics including the COVID-19 pandemic. We consider an initial period because the growth of the epidemic in this period typically happens before any public health intervention, such as contact tracing, preemptive quarantining, lockdown etc. and therefore represents the innate speed of the spread of the virus in the network, and depends only on the network structure and *β*. Using the data available in [17], we calculated this quantity for COVID-19 for different political units (country/region or province/state/dependency). We limited the analysis to political units that recorded at least 40 cases within the early stages of spread of COVID-19 (i.e., within first three months up to April 20, 2020) of the pandemic. There are 142 such countries/regions and 116 province/state/dependencies (Fig 2a). For each such political unit we considered *t*_0_ to be the time at which 40 cases are recorded in the unit, we consider that local community transmissions begin at *t*_0_; we consider *t* to be 3 weeks (21 days) from *t*_0_. We found that the growth rates in all these political units range from 0 to 0.31, with a median of 0.12 (Fig 2a). The range of *β* that we consider provides initial epidemic growth rates, in the diverse contact networks we consider, in a range that subsumes the realistic range [0, 0.31].

## Practical Aspects of Contact Tracing and Testing

We consider various attributes that affect the efficacy of contact tracing, involving variations of false negative rates, false positive rates, test result turnaround times, starting times of contact tracing, and level of cooperation with contact tracing and testing. We first set a default scenario and then consider a variety of environments departing from the choices in the default scenario based on estimates available in the literature. We first consider attributes of the testing equipment and logistics. Test results may be inaccurate, suffering from *false-negatives* and *false-positives*. A review [18] of 34 studies based on 12, 057 confirmed patients showed that false-negative rates ranged from 1.8 to 58%, with a median of 11%. We thus set the median 11% false-negative rate as default, but consider both the lowest and highest end-points of the reported range, though note that 58% is unrealistically high for the test-result to be meaningful. As for false positives, studies assessing a total of 119 South Korean laboratories [19, 20] and 52 Austrian laboratories [21] did not report false positive results, and a study evaluating 365 laboratories in 36 countries reported a false positive rate of 0.7% [22]. We set the 0% false-positive rate as default, but also consider 0.7% rate. Next, note that there is usually a delay between when a test is conducted and its result is obtained, this delay is known as the turnaround time. According to CDC [23], the turnaround times for 1) most nucleic acid amplification tests (NAATs), such as RT-PCR, vary between 1-3 days, 2) point-of-care tests are 15-45 minutes. We set default value of the turnaround time as 1 day, but also consider 3 days.

Public health authorities in different political units may decide to start contact tracing when the infection level in the target populace crosses a certain threshold. We consider that contact tracing is initiated when the first individual tests positive as the default option. This is in accordance with the observations of the leading practitioners of contact tracing programs who recognize that contact tracing should start as soon as the first case is diagnosed. Once the outbreak spreads, the logistical challenges associated with contact tracing multiply because of the sheer volume of the contacts that need to be traced [24]. Also, the only countries to have successfully contained the outbreak through contact tracing (i.e., before pharmaceutical preventives became available), namely South Korea, Japan, Vietnam, started the process early [6]. In order to understand the impact of the delayed initiations, we also consider the cases, e.g., 6 months and a year from when the outbreak is recorded. Using the data available in [17,25], we calculated the percentage of cumulative confirmed cases in different political units (186 countries and 137 states/provinces/dependencies) at the end of six months and a year from the date the datasets were recorded. The median of the percentages is 0.1% for 6-months delay and 1.1% for a year delay. Accordingly, we also consider scenarios in which contact tracing is initiated when the percentage of cumulative infections reaches 0.1% and 1.1%.

Finally, we assume full cooperation from the target populace as the default setting, i.e., every individual tests and quarantines as instructed by his local public health authority and reveals his contacts to them. But, we also consider scenarios in which cooperativity is less universal, and describe the forms and levels of non-cooperativity in the next section.

## Results

We quantify the costs and benefit of contact tracing over a course of 6 months (180 days) starting from the day after contact tracing is initiated. The total number of infections, the total number of tests, and the total sum of days of quarantine for the entire population over the period are averaged over 150 simulation runs, excluding those in which fewer than 40 individuals are infected within the first 3 months (90 days). By comparing the mean values of these results for different number of hops, we evaluate cost-benefit tradeoffs of multi-hop contact tracing scheme.

*Cost-benefit metrics –* We first define key cost-benefit metrics that are utilized throughout the evaluations. Recall that the *benefit* is defined as the percentage of reduction in the number of infections over the period of study compared to when no contact tracing was performed. There are two different kinds of *costs*: 1) tests and 2) quarantine. *Testing cost* is defined as the total number of tests over the period of consideration. We calculate the *quarantine cost* as the number of individuals quarantined on each day summed over all days in the period under consideration. This equals the number of days each individual is quarantined added over all individuals. We define *relative benefit* and *relative costs* to quantify the incremental benefits and costs multi-hop contact tracing provides/incurs as compared to 1-hop contact tracing. The *relative benefit for k-hop, k* > 1, is defined as the difference between the benefits provided by *k*-hop and 1-hop, i.e., benefit for *k*-hop - benefit for 1-hop. The *relative costs for k-hop, k* > 1, is defined as the ratio of cost difference between multi-hop and 1-hop to cost for 1-hop contact tracing, i.e., 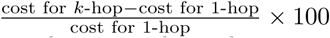, which represents how much more or less cost is required compared to 1-hop contact tracing.

First, we study the benefits and costs for multi-hop contact tracing over single hop (i.e., traditional) contact tracing. We show that the cost-benefit tradeoffs for multi-hop contact tracing can be classified into three phases, each corresponding to a different range of the growth rates; as the growth rate transitions into different ranges, sharp phase transitions are often observed. The range of growth rate we consider is [0, 0.31], the range in which those of the large number of political units we examined lie (Table S2 in SI Appendix). The classifications of the phases turn out to be robust to variations of false negative rates, false positive rates, test result turnaround times, delays in starting contact tracing, and even some form of non-cooperation such as when the non-cooperative individuals resume their activities from the date of the negative results discounting the possibility of false negatives. On the other hand, the classifications are significantly affected by the most debilitating forms of non-cooperation in which non-cooperative individuals neither disclose their contacts nor comply with public health guidelines on quarantining and testing.

Subsequently, we reveal that the further benefit for adding another hop beyond 1-hop tends to diminish progressively, thus a saturation phenomenon is observed. Accordingly, we investigate the highest number of hops (saturation point) that can lead to a non-negligible further reduction in the number of infections. We show that the saturation point increases as the growth rate increases and the environment becomes less conducive toward containing the disease, and is confined to 1, 2, and 3 hops in most cases.

## Single-hop vs Multi-hop Contact Tracing - A cost-benefit perspective

### Default Setting

We first evaluate the cost-benefit tradeoffs of multi-hop contact tracing under the default scenario for the synthetic networks. Fig 1c represents different networks and parameter combinations as points on a plot with growth rate as the horizontal axis and benefit as the vertical axis. This figure shows that despite the collective impact of various factors (types of contact networks, mean number of contacts per individuals, and transmission probability), the magnitude of the benefits provided by contact tracing can be characterized in terms of the growth rates. For the combinations in which the growth rate is low, 1-hop contact tracing alone can sufficiently contain the virus, and 2-hop and 3-hop contact tracing does not provide noticeable further benefit in terms of reduction in the outbreak size. Specifically, when growth rate is less than or equal to 0.105, 1-hop contact tracing reduces the outbreak size by 85.0 - 99.9% (median 97.8%). Next, a sharp *phase transition* is exhibited once the growth rate crosses a threshold value, that is, the benefit that 1-hop provides dramatically decreases, and multi-hop contact tracing offers substantial further benefit. Specifically, when the growth rate exceeds 0.105, 1-hop contact tracing reduces the outbreak size by 5.6 - 74.5% (median 28.1%), while 2-hop and 3-hop contact tracing respectively reduce the outbreak size by 54.5 - 99.92% (median 97.6%) and 84.6 - 99.96% (median 99.8%). This suggests that, as the virus spreads faster, traditional contact tracing becomes less than adequate. This is because in the time that elapses between when an individual becomes infectious and I is quarantined through 1-hop contact tracing, the disease spreads from I through a chain of several hops, i.e., the contacts I infects infect their contacts and so on. In this case, as shown in Fig 1a, preemptively tracing and quarantining multi-hop contacts can help tracing catch up with the speed of virus spread faster than 1-hop contact tracing.

We observe that 3-hop contact tracing can reduce the outbreak by 84.6 - 99.98% (median 99.8%) over the entire range of growth rates, as shown in Fig 1c. Hence, we quantify the costs and benefit of 3-hop contact tracing in comparison with 1-hop contact tracing. Fig 3 reveals that the relative benefit and relative cost for 3-hop contact tracing follow three *phases*:

**Fig 3.**
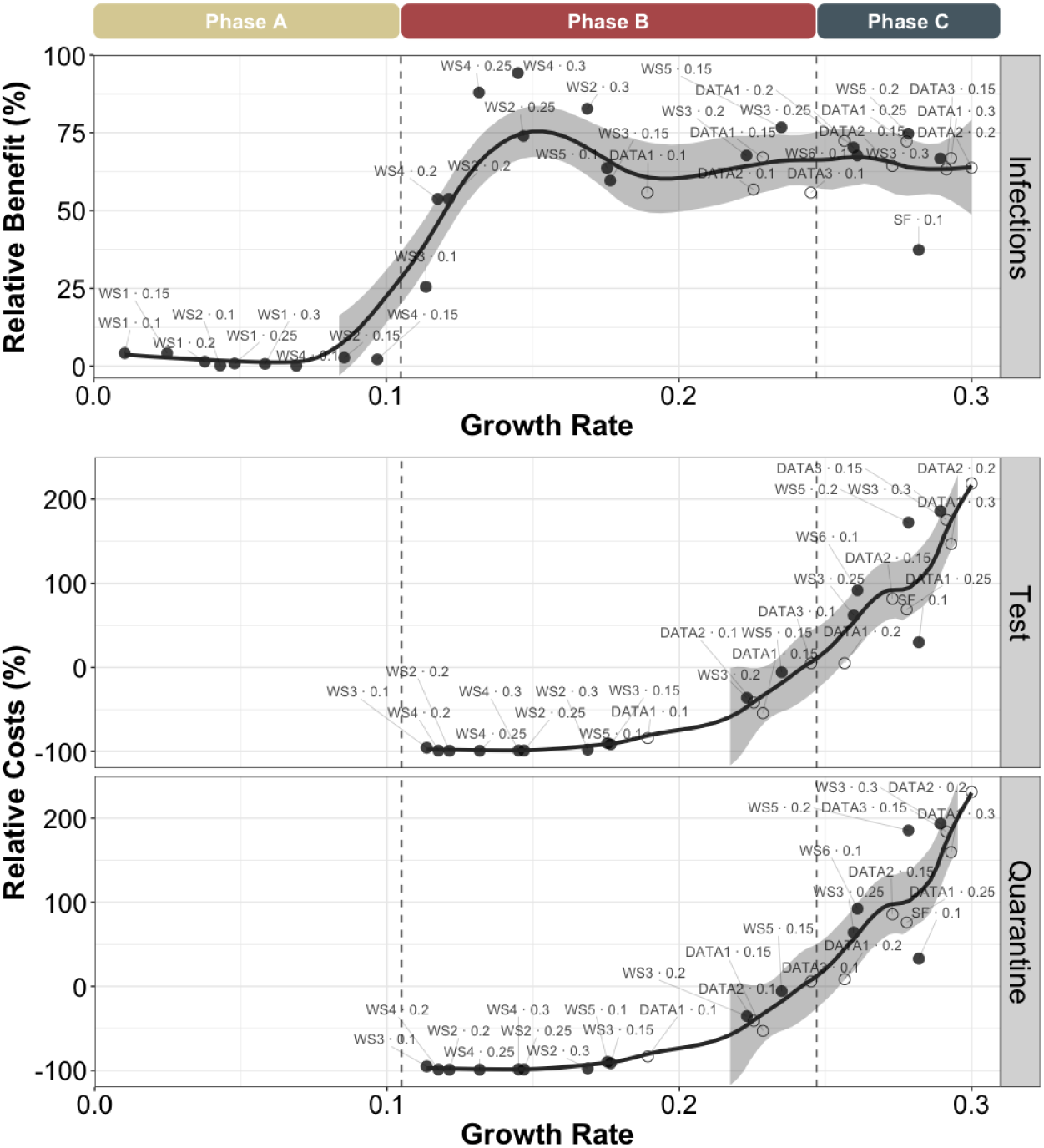
Relative benefit and relative costs. Relative benefit (the plot on top) and relative costs (the two plots on the bottom) are shown for 3-hop contact tracing under the default setting as a function of the growth rate. One can observe that the cost-benefit tradeoffs can be classified into three phases, depending on the value of the growth rate. Each point represents a combination of contact patterns and transmission probabilities and corresponds to a growth rate on the x-axis. For example, WS2. 0.25 represents that the contact pattern of a WS2 network (among the networks listed in Table 1) and the transmission probability is 0.25 and corresponds to the growth rate value of 74.0. The relative benefit and relative costs for the data-driven networks (shown in open circles) behave similarly to those observed at the same growth rates in synthetic networks (shown in filled circles). The solid lines correspond to the LOESS smoother with a span value of 0.3 and the shadings represent the 95% confidence interval around the smoother line. We determine the boundary between phases A and B as follows: compute the median value between the highest growth rate in group A and the lowest growth rate in group B for the synthetic networks and denote this value as the boundary. The boundary between phases B and C is defined similarly. The boundaries are depicted by lines.

- Phase A: In this phase, the relative benefit, as compared to single-hop contact tracing, is small (≤ 20%). Here, this corresponds to the regime of low growth rates (i.e., ≤ 0.105).
- Phase B: In this phase, the relative benefit increases substantially as compared to 1-hop while fewer total tests and fewer total sum of days of quarantine are needed for the entire population (relative benefit *>* 20% & relative costs ≤ 0%). Here, this corresponds to the regime of intermediate growth rates (between 0.105 and 0.247).
- Phase C: In this phase, multi-hop still provides a significant relative benefit, but requires greater costs compared to 1-hop tracing (relative benefit > 20% & relative costs > 0%). Here, this corresponds to high growth rates (larger than 0.247).

Multi-hop contact tracing may incur higher costs than 1-hop contact tracing because it traces up to more hops even from the same number of confirmed cases. However, this can more rapidly mitigate the spread of virus as compared to 1-hop contact tracing through faster identification and quarantine of multi-hop contacts of infected individuals, thus fewer individuals need tests with passage of time. In phase B, the latter phenomenon dominates, in phase C the former.

The above-mentioned simulation results on synthetic networks suggest that the cost-benefit tradeoff can be classified into three phases, depending on the value of the growth rate. We now verify this phenomenon on data-driven network. Recall that the parameter *r* represents the percentage of contacts between individuals of different villages. For different values of the parameter *r*, the growth rates for the networks fall into the regimes of intermediate-high. The relative benefit and relative costs behave similarly to those observed at the same growth rates in synthetic networks (see, e.g., the points that correspond to data-driven networks in Fig 3). Thus, the tradeoffs for the data-driven networks are consistent with those observed in the synthetic networks.

Next we study the role of different values of attributes, involving variations of false negative rates, false positive rates, turnaround times, starting times of contact tracing, and cooperativity. To this end, we revert to our synthetic networks and classify phases A, B, and C using the same criteria as in the default setting. For almost all variations of the attributes above (except for a debilitating form of non-cooperations that we will discuss), the cost-benefit tradeoff for multi-hop contact tracing can be still classified into three phases, with sharp transitions between them, depending on the value of the growth rate. In this respect, the classifications of the phases remains largely stable to variation of the above-mentioned attributes; the growth rate corresponding to the boundaries between phases tend to remain the same or shift to the left, as the environments become more challenging (Fig 4). The only exception is when we consider variation of the most debilitating form of non-cooperations (non-cooperative individuals not revealing their contacts, not testing nor quarantining). In this case, the cost-benefit tradeoff is significantly altered because refusal to test and reveal contacts limit tracing and the contact network as known to tracers becomes highly fragmented and sparse.

**Fig 4.**
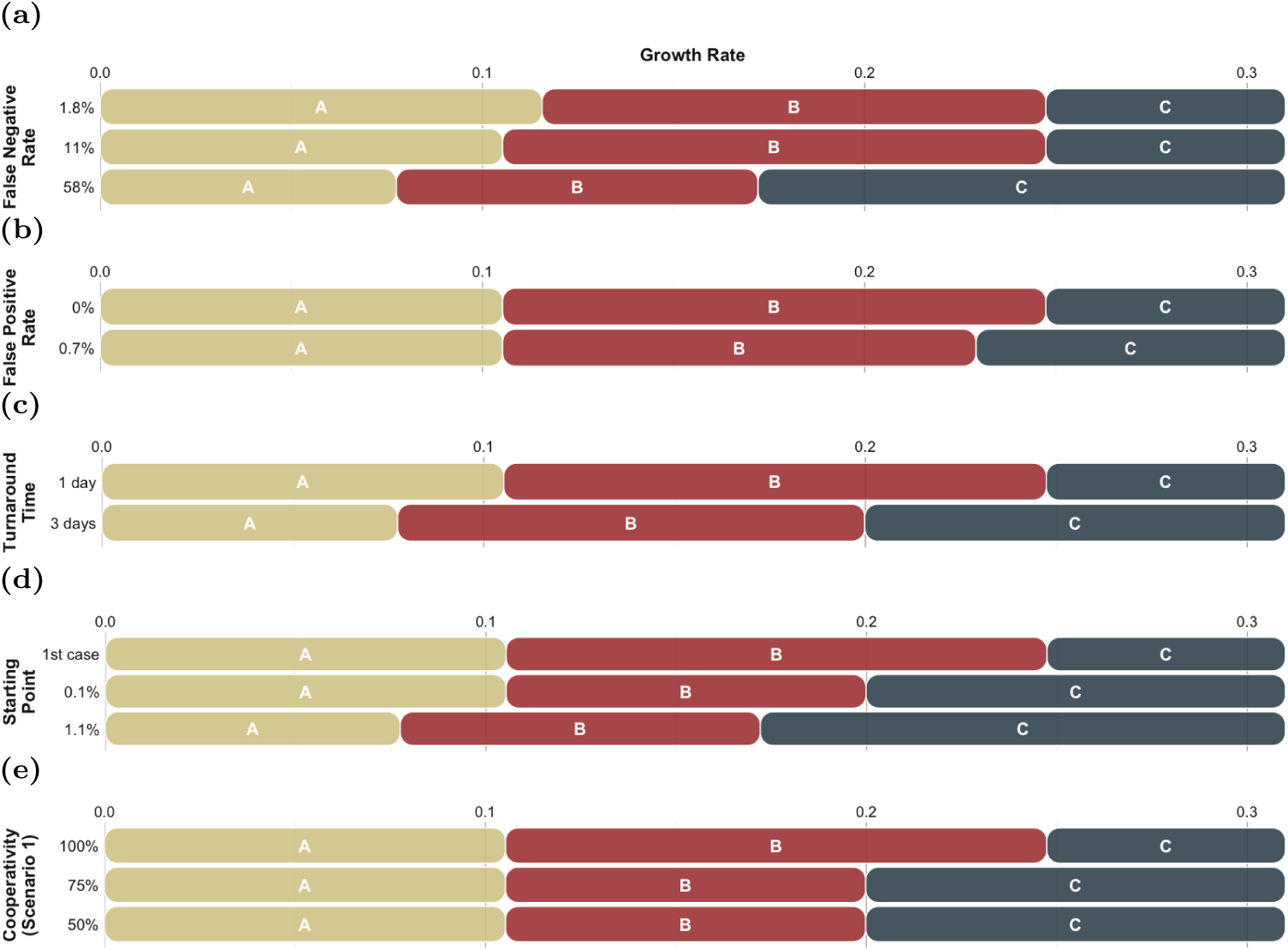
Classifications of the phases for variations of practical aspects of contact tracing and testing. Classifications of the phases for variations of (a) false negative rate, (b) false positive rate, (c) test result turnaround time, (d) starting points of contact tracing, and (e) level of cooperation with contact tracing and testing (Scenario 1). Different capital letters inside each bar represent different phases. Even for the wide variations of the attributes, the cost-benefit tradeoff for multi-hop contact tracing can be still classified into three phases, with transitions between them, depending on the magnitude of the growth rate value. In this respect, the classifications of the phases remains largely stable to variation of the above-mentioned attributes; the growth rate corresponding to the boundaries between phases tend to remain the same or shift to the left, as the environments become more challenging.

### Properties of Tests

False negatives prevent tracing the contacts from the individuals who test negative but are infected in reality. This can reduce the benefits of contact tracing. This also may reduce costs as fewer individuals need be quarantined and tested, but on the other hand may also increase costs as larger outbreaks may eventually require higher number of tests and quarantines. We assess the impact of these two opposing factors. Fig 4a shows that the growth rates corresponding to the boundaries between phases remain similar or shift to the left, as the false negative rate increases. The boundary between Phase A and B is 0.116 for 1.8% false-negative rate, 0.105 for 11% rate, and 0.077 for 58% rate; the boundary between Phase B and C is 0.247 for 1.8% rate, 0.247 for 11% rate, and 0.172 for 58% rate.

False positives do not reduce the benefit of the contact tracing strategies, but may incur higher costs particularly for greater values of the growth rate. Fig 4b shows that the growth rates corresponding to the boundaries between phases remain similar or shift to the left, as the false positive rate increases. The boundary between Phase A and B is 0.105 for 0% false-positive rate, and 0.105 for 0.7% rate; the boundary between Phase B and C is 0.247 for 0% rate, and 0.229 for 0.7% rate.

### Test Result Turnaround Time

Longer test result turnaround time causes delays in contact tracing. Fig 4c shows that the trend with regard to the growth rate is nonetheless similar to what was explained before.

### Starting Point of Contact Tracing

We now assess how the cost-benefit tradeoff is affected if public health authorities do not initiate contact tracing right at the onset of the epidemic. We allow the virus to spread on contact networks without any intervention. We consider different scenarios: 1) start contact tracing when the first individual tests positive, 2) start contact tracing when the cumulative infection percentage crosses a designated threshold. In the latter case, we consider two different thresholds, 0.1% and 1.1%. Fig 4d shows that the trend with regard to the growth rate is nonetheless similar to what was explained before. We calculated averages over 150 simulation runs in which this threshold is crossed within one year, and consider only the networks and parameter values for which this threshold is crossed for at least 1 simulation run among the first 150 runs.

### Level of Cooperation with Contact Tracing and Testing

The extent of cooperation of individuals with public health guidance plays an important role in preventing the spread of the virus. Non-cooperation can arise in different forms: individuals not revealing their contacts, not testing nor self-quarantining. Based on the default setting, we explore two different scenarios of limited cooperation:

- Scenario 1: Non-cooperative individuals resume their activities from the date of the negative results discounting the possibility of false negatives, and do not share their contacts during the period of advised quarantine.
- Scenario 2: Non-cooperative individuals do not get tested nor quarantine upon notification of exposure. They get tested and quarantine only if they develop symptoms and that too after a delay from symptom onset. They do not disclose their contacts either before or after getting tested.

Fig 4e shows that in Scenario 1, the trend with regard to the growth rate is nonetheless similar to what was explained before. However, in Scenario 2, the phases become much less clearly demarcated based on the value of the growth rate. This is because, in Scenario 2, as fewer individuals cooperate, fewer individuals reveal their contacts and get tested, thus very few individuals can be tested through multihop tracing and the contact network as known to tracers becomes severely fragmented and sparse. Even for 75% cooperativity, the trend with regard to the growth rate starts not to hold; for 50% cooperativity, the relative benefit decreases and becomes below 20% in many cases, and thus they belong to phase A (Fig 5).

**Fig 5.**
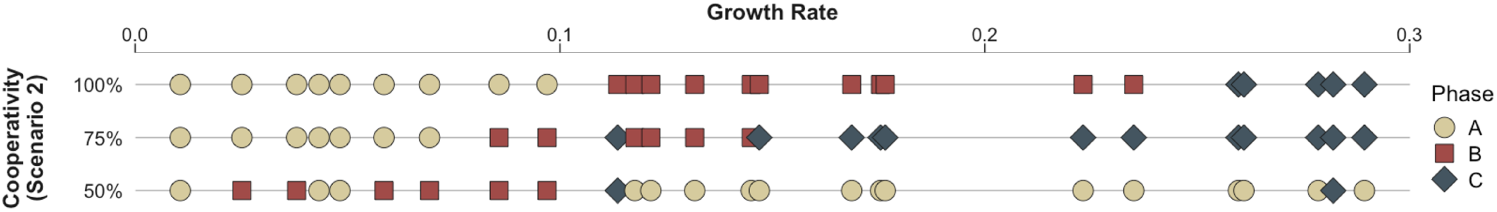
Classifications of the phases for variations of level of cooperation (Scenario 2). When this form of cooperativity is low, the cost-benefit tradeoff for multi-hop contact tracing is no longer classified by the growth rate. In particular, when 50% of individuals do not cooperate, multi-hop contract tracing process itself can be vigorously challenged especially by the individuals who do not reveal their contacts. Hence, most cases belong to phase A in which multi-hop contact tracing does not provide significant further benefit beyond what 1-hop provides.

## Diminishing Returns for Increasing Number of Hops

We have shown that the cost-benefit tradeoffs can be classified into three phases, with transitions between them, depending on the value of the growth rate, and can be substantially enhanced through the deployment of a natural multi-hop generalization through comparison between traditional (1-hop) and 3-hop contact tracing. In this section, we investigate the highest number of hops that can lead to a non-negligible further reduction in the number of infections. Our numerical computations reveal that the further benefit for adding another hop beyond 1-hop tends to diminish progressively, thus a *saturation phenomenon* in which the benefit increases only marginally by increasing the number of hops beyond a certain point arises. Hence, increasing the number of hops beyond the hop number at which the saturation phenomenon is observed is not effective when it comes to reducing the number of infections. Our criteria is that the saturation point becomes *k*-hop when further benefit provided by (*k* + 1)-hop over the previous *k*-hop is less than 10% (i.e., difference between benefits of (*k* + 1)-hop and *k*-hop is less than 10%).

We pooled all the results across variations of practical aspects of contact tracing and testing (involving variations of false negative rates, false positive rates, turnaround times, starting times of contact tracing, and cooperativity (Scenario 1)), and observed that there are broad trends, with regard to the saturation points, across variations of the environments above. Specifically, the saturation point becomes greater as the growth rate increases (Fig 6) or the environments become less conducive toward containing the disease. Even for wide variations of the attributes above, marginal histogram along the y-axis in Fig 6 shows that the saturation point is confined to 1, 2, and 3 hops in most cases (89% of all instances), while the saturation points of 4 and 5 hops largely arises for very limited conditions such as higher growth rates in more challenging environments (11% of all instances).

**Fig 6.**
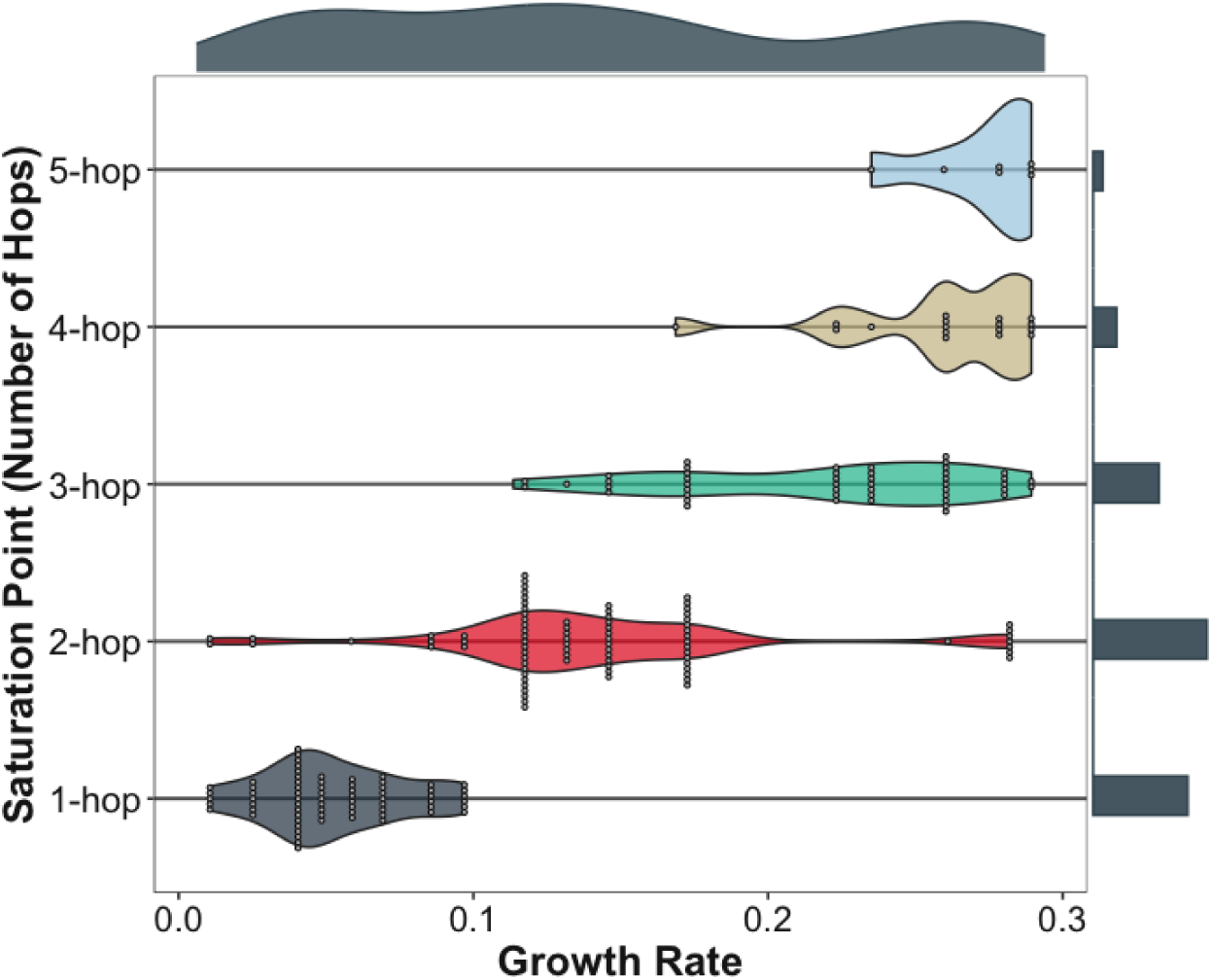
Saturation point. Violin plots show the distribution of growth rate across discrete saturation points, based on the pooled data across the environments (involving variations of false negative rates, false positive rates, turnaround times, starting times of contact tracing, and cooperativity (Scenario 1)). We use the same synthetic networks and values of transmission probability that are used in Fig 4. Each point corresponds to a certain contact pattern, a certain transmission probability, and a certain environment, which determine the growth rate (x-axis). Marginal distributions are added to the margin of each axis. The saturation point becomes greater as the growth rate increases; the instances of the saturation point being 1 hop is mostly concentrated in low growth rate region, 2 hops in intermediate region, and 3 hops and more in high region. Furthermore, marginal histogram along the y-axis shows that the saturation point is confined to 1, 2, and 3 hops in 89% of all instances, while the saturation point becomes 4 or 5 hops in 11% of instances corresponding to higher growth rates in more challenging environments.

In addition to the saturation points, we seek to reveal some broad trends with regard to the choice of the number of hops, recurring across various environments, taking into account both further benefit and cost incurred by each hop over the previous. Our simulations show that the broad trends resemble the trends with regard to the saturation points; as growth rate increases or environments becomes more challenging, the cost-benefit tradeoffs propel us towards choosing higher number of hops. Refer to SI Appendix for details on specific numbers, analysis, and criteria for choosing the number of hops, in which we present the further benefits and costs up to 5 hops for different parameter combinations.

## Discussion

Contact tracing has been deployed during the first year of the pandemic in many countries, but very few of those countries have successfully contained the pandemic before the advent of pharmaceutical interventions. Vietnam is one of the few success stories in successful containment of the outbreak in early stage of pandemic, and it is also the only country to have incorporated multi-hop contact tracing in its containment program. Since contact tracing (and quarantine) remains one of the few available mechanisms to contain the outbreak and prevent a pandemic during the early phases of any epidemic, an independent influence of the multi-hop contact tracing policies in the absence of any public health intervention needs to be comprehensively and systemically investigated. It is crucial to examine when and why traditional contact tracing is not sufficient to contain the virus and whether the multi-hop contact tracing can enhance the cost-benefit tradeoffs in such circumstances.

In this work, we embarked on an investigation of multi-hop contact tracing considering a diverse set of large-scale contact networks, spanning synthetic networks of various families and design choices and those obtained from real-world interaction data. We now summarize and position our results. First, our findings confirm the intuition that multi-hop contact tracing reduces the spread of the infection. Next, our findings however go beyond, by revealing patterns that can not be intuited a priori. We reveal that multi-hop contact tracing has the potential to reduce the outbreak to a much smaller size as compared to conventional contact tracing (i.e., 1-hop contact tracing), *even at lower costs* than the conventional contact tracing. We also show that the cost-benefit tradeoffs for multi-hop contact tracing can be classified into three phases, with sharp transitions between the phases, and each phase corresponds to a different range of the initial epidemic growth rates. When the growth rates are low, multi-hop Nbecomes redundant as single-hop contains the outbreak adequately. For higher growth rates, multi-hop substantially reduces the outbreak size, incurring 1) substantially lower quarantining and testing costs as compared to single-hop in the intermediate growth rate region 2) considerably higher costs in the high growth rate region. Furthermore, the classifications of the phases turn out to be robust to wide variations of almost all the practical aspects of contact tracing and testing.

Third, we show that the further benefit of adding another hop beyond 1-hop tends to diminish progressively, thus saturation phenomenon arises. While it is intuitive that there would be a saturation phenomenon, the impact of the growth rates on the saturation point in different ambiences can not be inferred without the quantitative investigation. As growth rate increases or the contact tracing and testing ambience becomes more challenging, the saturation point becomes greater and the cost-benefit tradeoffs propel us towards choosing higher number of hops. We calculate the growth rates in a large number of political units from publicly available pandemic data; our calculations show that these growth rates span all three ranges. In particular, therefore, multi-hop contact tracing substantially reduces the outbreak size and lowers overall costs for a large number of realistic values of growth rates.

Multi-hop contact tracing has been subject to limited rigorous investigation thus far. To our knowledge, the only other work to investigate this concept has been [26]. Our work is complementary to [26] which used real-world social network data of 468 individuals and considered tracing and quarantining (without testing) both primary and secondary contacts of those who test positive. [26] found that quarantining secondary contacts decreases the cumulative infection count compared to quarantining only the primary contacts, but also requires substantially higher number of quarantines. Next, they focused on reducing the number of quarantines through 1) social distancing and 2) testing. When individuals are tested, those who test negative are released from quarantine right after the results are obtained; this reduces the quarantine periods but increases the outbreak. The authors acknowledge that it is unclear if their results would hold for networks with larger size. Results may become artifacts of network size for multi-hop contact tracing because the length of contact chains may be limited by network size when the size is small. We investigate multi-hop contact tracing involving a combination of quarantining and testing for *k* hops, where *k* can be 2, 3, 4, 5 etc. over large networks comprising of up to 100, 000 individuals, and consider a large number of instances from both synthetic networks corresponding to various families and parameter choices and networks obtained from contact data. We use tests to further trace contacts rather than to release those traced early from quarantining and evaluate both testing and quarantining costs. As mentioned in the previous paragraph, we show that the cost-benefit tradeoffs for different number of hops (1, 2, 3, 4, 5, …) can be very different depending on growth rate and venue of the tests, and the tradeoffs for different test venues can be characterized by only one parameter vis a vis the network topology, that is the growth rate. In particular the result [26] reports as to the comparison between 1 and 2 hops for “quarantine only” corresponds to what we observe throughout the high growth rate range for our simulations. When social distancing is additionally incorporated, the growth rate decreases; their finding in this case is consistent with the phenomenon we observe for the intermediate growth rate range. Thus, our investigation positions their findings as parts of a broader trend. This is in addition to revealing the phase-transition patterns for cost-benefit tradeoffs and identifying the hop choices for different ranges of growth rates, different testing ambiences and a diverse class of larger networks.

We now discuss limitations of multi-hop contact tracing in the current context and how to circumvent the limitations in order to prevent a future epidemic from becoming a pandemic. First, the benefits of contact tracing, both single hop and multi-hop, considerably decrease if a non-negligible percentage of the society do not reveal their contacts, do not test, and do not quarantine when asked to. Cooperation with health authorities varies across the world: while a high degree of cooperation was witnessed in South Korea and Taiwan which had suffered from large scale epidemics in the last twenty years [27, 28], cooperation was lower in Europe and US [29], both of which experienced a large scale epidemic about a century ago (the 1918 flu). Learning from the experience of this pandemic, public awareness campaigns need to be pursued to elicit cooperation with health authorities. Multi-hop may provide an important advantage to ensure cooperation in that it can contain the outbreak faster which may incentivize full cooperation for a short duration, whereas cooperation may wane due to pandemic-fatigue as time progresses.

We next describe the generalization of our framework for an investigation for other infectious diseases. Each infectious disease differs from the other in two aspects such as stages of the disease evolution and parameters for the disease. The investigation on cost-benefit tradeoff of multi-hop contact tracing for an arbitrary infectious disease can be extended by appropriately choosing the stages and parameters for the disease based on our framework. Almost all infectious diseases include susceptible, recovered and dead stages, the choice of other states allow us to cater a specific disease in question. As for COVID-19, latent, pre-symptomatic, symptomatic, and asymptomatic are the additional stages. Let us consider smallpox as an example of another infectious disease. All individuals infected with smallpox develop symptoms (fever and rash), thus latent and symptomatic stages can be added and the symptomatic stage can be further subdivided into fever, early rash, and late rash stages. Smallpox does not have asymptomatic carriers, so the asymptomatic stage can be omitted [30]. The different stages and parameters for the disease in question alter the epidemic growth rate. Question that remains is that if the observed patterns regarding the cost-benefit tradeoff for multi-hop contact tracing, namely the phase classifications, the sharp phase transitions, the saturation phenomenon, extends to other infectious diseases. Investigating multi-hop contact tracing for other infectious diseases based on our framework constitutes an imperative direction for future research towards building a knowledge-base for containing future epidemics before they become pandemics and repeat the enormous toll that COVID-19 imposed.

We now enumerate some additional directions for further research. We have assumed that the traced *k*-hop contacts of an individual (say I) who tests positive can start their quarantine within a day of I testing positive, though they test after some delay. But this is not in general possible unless I downloads the contact tracing app either before or at least right after testing positive [31]. Next, depending on classifiers such as duration, environment (indoor or outdoor), usage of protective equipment, observance of personal hygiene, different contacts may pass on infection with different probabilities. Assuming that such a probability is identical for all contacts with same infectious categories, which is what we did, is equivalent to considering an average over all contacts. Explicitly investigating the impact of 1) delays in starting quarantining and 2) non-uniform transmission probabilities constitute directions for future research.

## Methods

### Compartmental model of virus transmission

Compartmental models have been widely used in studies on virus spread [32,33]. We use a discrete time compartmental disease model to model the progression of COVID-19 where the transition from each compartment to the next happens after a random amount of time with a geometric distribution. Different stages of the disease are shown in Fig 2b. The model consists of the following stages: Susceptible (*S*), Presymptomatic-Latent (*I*_*p*_-*L*), Presymptomatic (*I*_*p*_), Symptomatic (*I*_*s*_), Ready-to-Test (*RT*), Asymptomatic-Latent (*I*_*a*_-*L*), Asymptomatic (*I*_*a*_), Recovered (*R*), and Dead (*D*). Only symptomatic individuals show symptoms, while presymptomatic, symptomatic and asymptomatic individuals can infect others. When a susceptible individual comes into contact with an infectious individual, the susceptible is infected with transmission probability *β*.

Once an individual is infected he becomes contagious after a geometrically distributed latency time, whose expectation depends on whether he will develop symptoms at some point or otherwise. Following the nomenclature in compartmental models already utilized for COVID-19, we assume that an infected individual becomes asymptomatic-latent (with probability *p*_*a*_) or presymptomatic-latent (with probability 1 − *p*_*a*_) and those in this latency period have a negative test (the tests do not detect the presence of COVID-19). The asymptomatic-latent (*I*_*a*_-*L*) individuals never develop symptoms, do not infect others for a mean latency duration of 1*/λ*, and subsequently become contagious, at which stage we call them asymptomatic or *I*_*a*_ for simplicity. An asymptomatic individual remains contagious for a geometrically distributed random duration with mean 1*/r*_*a*_, after which the individual recovers. We now consider the other compartment an individual enters after infection, the presymptomatic-latent compartment. A presymptomatic-latent individual, say *B*, becomes contagious after a mean latency period of 1*/λ*, at which point we call *B* presymptomatic or *I*_*p*_. *B* remains presymptomatic for a geometrically distributed duration with mean 1*/α*; after this duration *B* develops symptoms and is called symptomatic. A symptomatic individual *B* continues to infect contacts until *B* opts for testing (*RT*). The duration for which a symptomatic individual infects others is geometrically distributed with mean 1*/w*. Once this duration ends, the patient quarantines and does not infect others. The patient ultimately dies (*D*) with probability *p*_*d*_, or recovers (*R*) with probability 1 − *p*_*d*_, after a geometrically distributed duration whose mean is 1*/r*_*s*_. We do not consider that individuals can be reinfected. In all the networks, we consider that initially all but three individuals are susceptible, among the three there is one presymptomatic, one symptomatic and one asymptomatic. Refer to Table S1 in SI Appendix for the parameter values we choose.

### Synthetic networks

We consider two classes of synthetic networks: 1) Watts-Strogatz networks [10] and 2) scale-free networks [34]. Each network we consider has *N* = 100, 000 nodes. The Watts-Strogatz networks have average degrees of ⟨*k*⟩ = 4, 8, that is, 200, 000, 400, 000 edges. They are generated following a variant of the original Watts-Strogatz model. Based on a ring of *N* nodes, each node is connected to *k* nearest neighbors by undirected edges. Subsequently, each end point of each edge is rewired to a uniformly randomly chosen node over the entire ring with rewiring probability of *p*, avoiding link duplication (i.e., multiple edges between the same pair of nodes) and self-loops. The scale-free network topologies are generated by Barabási-Albert model where new nodes are added at each time step with *m* links that connect to existing nodes with a probability that is proportional to the degree of the existing nodes [34]; we set *m* = 2 to generate the network. The resulting network consists of 199, 997 edges, thus average degree of a node is ⟨*k*⟩ = 3.99994.

### Data-driven network

We use the publicly-available network data covering a wide range of interactions among individuals collected by survey in each of 75 villages located in Karnataka, India [15]. The surveys includes interaction information such as names of those who visit the respondents’ homes, those with whom the respondents go to pray, etc. The network consist of a total of 69, 441 individuals and 294, 945 interactions among them. However, the dataset only contains information on interactions between individuals within each village. Since the population size of each village is relatively small population (354 to 1775 individuals for each village), we introduce interaction between individuals in different villages through degree-preserving rewiring [35,36]. We first randomly select two villages and select a random edge within each cluster, and then swap the two edges to reach across the pair of villages. The process is repeated until the percentage of edges that are rewired among the total number of edges becomes *r*%, and *r* is called the mixing parameter [36]. The degree-preserving rewiring preserves the degree of all the nodes in the network regardless of the parameter *r*, but it changes the frequency of inter-village interactions and network properties. We generated three different networks with the mixing parameters *r* = 1, 3, 5. As the parameter *r* increases from 1 to 5, the diameter, average path length, and clustering coefficient monotonically decrease (refer to Table 1).

## Data Availability

Relevant data are within the manuscript and Supporting Information. Contact networks generated during this study are available from the repository https://github.com/jungyeol-kim/contact-tracing.

## Supporting information

**S1 Appendix. Contains the following:** Section 1. Digital Contact Tracing; Section 2. Choice of the Number of Hops; Table S1. Values of Disease Parameters; Table S2. Real-world Initial Epidemic Growth Rates; Table S3. Further Benefit/Cost Across Different Number of Hops (Data-driven Network); Table S4. Further Benefit/Cost Across Different Number of Hops In *Low* Growth Rates Region (Synthetic Networks); Table S5. Further Benefit/Cost Across Different Number of Hops In *Intermediate* Growth Rates Region (Synthetic Networks); Table S6. Further Benefit/Cost Across Different Number of Hops in *High* Growth Rates Region (Synthetic Networks).

## S1 Appendix

### Digital Contact Tracing

Apps have already been deployed by states in US and other countries that can use technology developed by Apple and Google and anonymous Bluetooth signals to digitally trace and notify the 1-hop contacts of COVID-19 patients who test positive (e.g., COVID Alert Pennsylvania, COVID Green in Ireland, Delaware COVID-19 tracing app [1]). These apps can trace and notify 1-hop contacts over last 14 days even if patients download the apps after they test positive. Thus, a recursive utilization of this app can be used to trace *k*-hop contacts, that is, if 1-hop contacts download such apps after notification of their exposure, tracing their 1-hop contacts over the previous 14 days will provide the 2-hop contacts of the patient who tested positive over the last 14 days and so on. The notified individuals receive an alert to check the app, which provides instructions and information from state health officials about seeking medical help, staying at home, and quarantining. To ensure privacy, the app shields the identity of the person who tested positive from people receiving a notification, and vice versa. It does not store location data, personal information, or the identities of individuals who were possibly exposed and keeps the data anonymous [1].

### Choice of the Number of Hops

Our numerical computations reveal some broad trends with regard to the choice of the number of hops, recurring across various environments, taking into account both further benefit and cost incurred by each hop over the previous. We first define two additional concepts. The *further benefit of hop k* is defined as additional benefit provided by increasing hops by one, i.e., difference between benefits of *k*-hop and (*k* − 1)-hop. The *further cost of hop k* is defined as the additional relative cost incurred by increasing hops by one, i.e., difference between relative costs of *k*-hop and (*k* − 1)-hop. While criteria for choosing the number of hops may be subjective, for ease of exposition, we rule out higher number of hops if they do not result in 10% or greater further benefits. Recall that we chose a 20% difference in relative benefit for 3 hops as the criteria for classifying instances of topologies and parameters into phase A vs phases B, C. The criteria for choice of hops is applied on a different quantity, further benefit of higher number of hops. Different values for the two markers, 10%, 20%, have been used because the goal for the demarcations are different: 1) a higher difference 20% is used to demarcate the instances in which multi-hop substantially lowers the outbreak size as compared to 1-hop (phases B, C) 2) higher hops are ruled out when they provide only marginal further benefit, that is, lower than 10%. Among the rest of the hops, we posit that the choice would be made based on additional costs incurred by each hop over the previous. Recall that, for the default setting, phases A, B, and C respectively correspond to growth rates of 0 - 0.105, 0.105 - 0.247, and 0.247 - 0.31. We refer to these fixed ranges as *low, intermediate* and *high* throughout the paper. We present broad trends observed in each region for a wide range of parameter choices, based on the criteria mentioned above:

- In the low growth rate range, 1-hop contact tracing alone frequently reduces the outbreak size by over 90%, and further benefit on increasing the number of hops beyond 1-hop is usually lower than 10%. Thus, 1-hop is the natural choice by our criteria. We observe deviation from this trend in some instances as the environments become more challenging. When the further benefits are higher for multi-hop contact tracing, 2-hop suffices in reducing the outbreak size and also attains lower cost as compared to 1-hop in almost every case.
- In the intermediate growth rate range, the benefit that 1-hop provides dramatically decrease as compared to the low growth rate range (sharp phase transition once a threshold value is crossed), and 2-hop offers substantial further benefit, even at a lower cost compared to 1-hop. Increasing the number of hops beyond 3-hop provide lower than 10% further benefit, except for a few instances in which cooperativity is low and false negative rates are excessively high. Thus, except for such challenging environments, the choice can be limited to 2 or 3 hops in this range per our criteria. Between 2 and 3 hops, the further benefit provided by 3 hops exceeds 10% only in some instances towards the higher end of the intermediate growth rate range. Thus, per our criteria, 2 hop constitutes a natural choice in most of the intermediate growth rate range. We observe divergence from this trend as the environments become more challenging, such as, excessively high false negative rates, and low cooperativity. Also, in general, the trends in the higher end of the intermediate growth rate range resemble those in the high growth rate region. Finally, wherever the further benefits attained by 3-hop exceeds 10%, the choice between 2 and 3 hop would be determined based on the additional cost incurred by 3 hop over 2 hop.
- In the high growth range, further benefit of 2-hop and 3-hop mostly exceed 10%, those of 4-hop also exceed 10% in several instances, those of 5-hop exceed 10% in very few instances. Usually, each hop fetches greater cost than the previous hop. This constitutes an important distinction with intermediate growth rate range. In the intermediate growth rate range 2-hop usually incurs lower cost than 1-hop except for very challenging environments and the few highest growth rates in the range, and in several of these 3-hop incurs lower cost than 2-hop. In the high growth rate range, additional benefits provided by 4 and 5-hop increase as the environments become more challenging, such as, increase in false negative rates and turnaround times and decrease in cooperativity (Scenario 1). The choice on the number of hops need to be made depending on the magnitude of the additional benefits and affordability of the additional costs incurred when hop count is increased (this is also the phenomenon observed in extreme scenarios and in growth rates at the higher end of the intermediate growth rate range).

Thus, the choice is mostly confined to 1, 2, 3 hops, with the need for considering 4 and 5 hops largely arises for limited conditions such as high growth rates and very challenging environments. Next, in general, the need for choosing a larger number of hops becomes greater as the growth rate increases or the environment becomes less conducive towards containing the disease. But, at the same time the further benefit for adding another hop beyond 1-hop tends to diminish progressively in almost every topology, just that, the decrease becomes less pronounced for higher growth rates and in more challenging environments.

### Default Setting

We now describe the simulation results in the default setting in greater detail. The further benefit incurred by adding another hop beyond 1-hop tends to diminish progressively. In a range of low growth rates, 1-hop contact tracing reduces the outbreak size by 85.0 - 99.9% (median 97.8%), and the further benefit provided by increasing the number of hops beyond 1-hop is less than 4.3%.

When the growth rate exceeds a certain threshold, we observe a sharp decrease in the benefit that 1-hop provides. Specifically, in a range of intermediate growth rates, the benefit provided by 1-hop is 5.6 - 74.5% with 30.3% as median. In all but two instances in this region, 2-hop adequately contain the outbreak (further benefit beyond what 1 hop provides is 25.5 - 93.9%, median 63.0%); increasing the number of hops beyond 2-hop provides only 2.4% or lower further benefit. The costs for 2-hop are even lower than those for 1-hop (further costs for 2-hop are negative). In the remaining two instances, which correspond to the highest growth rates in the intermediate growth rate range, 2-hop offers further benefit of 51.6% and 59.9% and incurs higher costs than 1-hop, simultaneously 3-hop also provides further benefit of 16.0% and 16.9% with less costs than both 1 and 2-hop (further cost of tests and quarantine for 3-hop is respectively − 57.8, − 32.9 and − 58.0, − 33.2). Also, increasing the number of hops beyond 3-hop provides only at most 1.8% further benefit in these instances. Thus, 3-hop emerges as the natural choice in these instances. Thus, in the intermediate growth rate range, 2-hop constitutes the natural choice, except for two instances at the high end of this range, for which 3-hop constitutes the natural choice.

In the high growth rate region, further benefits of 1-hop, 2-hop, 3-hop, 4-hop are 12.4-43%, median 23%, 34.8 - 56.5%, median 42.2%, 2.5 - 32.6%, median 27%, 0.3 - 9.8%, median 4.6%, respectively. Thus, starting 2 hops, the further benefits are progressively decreasing, but are considerable for up to 3 hops. Also, multi-hop contact tracing (up to 4 hops) incur greater costs than that incurred by the previous hop in almost every case; further cost of tests and quarantine for 3-hop is respectively up to 53.4 and 51.4, and further cost of tests and quarantine for 4-hop is respectively up to 100.9 and 95.7. By our criteria, the choice should be between 2 and 3 hops, depending entirely on the affordability of the additional costs incurred by 3 hop in each instance.

Thus, the natural choices that emerge are: 1) 1-hop throughout the low growth rate region 2) 2-hop throughout the intermediate growth rate region except for 2 with the highest growth rates in which 3-hop constitutes the natural choice 3) either 2 or 3 hops throughout the high growth rate region depending on the affordability of the additional costs incurred by 3 hops in each instance. Also, for each topology, starting from 2 hops, the further benefits constitute a non-increasing function of the number of hops, thus the principle of *diminishing return* holds.

The broad trends also recur in the data-driven networks. Recall that for different values of the parameter *r*, the growth rates for the networks are distributed in the second half of the intermediate range and in the high range. In the intermediate range, the benefit provided by 1-hop is 30.8 - 43.7% with 42.1% as median. 2-hop contact tracing provides further benefit of 47.6 - 57.7% median 53.6%. Increasing the number of hops beyond 2-hop provides 9.4% or lower further benefit. The costs for 2-hop are even comparable or lower than those for 1-hop (further costs for 2-hop are negative except in 1 instance). Thus 2-hop constitutes an appropriate choice in this region.

In the high range, further benefits of 1-hop, 2-hop, 3-hop, 4-hop are 10.9 - 29.7%, median 21.3%, 29.3 - 44.5%, median 36.4%, 22.5 - 38.4%, median 29.2%, 3.4 - 13.7%, median 6.0%, respectively. Starting 2-hop, the further benefits are mostly progressively decreasing. Also, the further costs of these hops are all positive, except some cases for 3-hop. By our criteria, the choice will be between 2 and 3 hops, except in one instance in which the further benefit for 4-hop exceeds 10% in which case 2, 3, and 4 hops should be considered. Thus, the phenomena previously observed in various types of synthetic networks also recur in the data-driven networks.

### Level of Cooperation with Contact Tracing and Testing: Scenario 1

The trends are same as the default scenario with the following differences. For 50% cooperativity, the behavior of the instances with the two highest growth rates in the intermediate growth rate range become identical to those in high growth rate region. Specifically, each hop incurs higher costs compared to the previous hop. The further benefit attained by 4-hop exceeds 10%, and in one of these two instances, the further benefit of even 5-hop exceeds 10%. Thus, in these two instances, the choice would be between 2, 3, and 4 hops or even 2, 3, 4, and 5 hops and will depend on the affordability of the additional costs of each hop.

### Properties of Tests

As noted earlier, false positives primarily increase the costs for greater number of hops in the high growth rate region; further cost of tests and quarantine for 3-hop is respectively up to 313.4 and 319.1 for 0.8% false positive rate. Otherwise, the trends are the same as that for the default scenario.

Under false negatives, the trends for 1.8% false negative rate is very similar to those for 11% (default), which means that the trends in general and the effectiveness of multi-hop contact tracing in particular are resilient to increases in the false negative rate in a realistic range. However, we notice actual divergence from the broad trends reported above only for the impractically high false negative rate, 58%, which we consider for the sake of completeness. We describe the divergences next.

Considering the low growth rate region, in two instances towards the upper end, 2-hop provides greater than 10% further benefits beyond what 1-hop provides (62.4, 70.5%) with less costs than 1-hop. The behavior of those instances with highest growth rates in the low growth rate region is similar to those in the intermediate growth rate region. Recall that in the earlier subsection of the Results section, we had noted that phase B starts at the higher end points of the low growth rate region for the same setting. The phenomena here is consistent with the earlier observation.

In the intermediate growth rate region, the further benefit of 5-hop is lower than 2.8% throughout. So, the choice ought to be between 2, 3, and 4 hops. 2-hop always, 3-hop mostly and 4-hop in some instances, provide greater than 10% further benefit. Also, except for 2 instances which have the highest growth rates, 3-hop attains lower cost compared to 2-hop which attains lower cost compared to 1-hop, 4-hop attains comparable or lower costs compared to 3-hop. Thus, 3 or 4 hops constitute the natural choices in most of these instances (wherever its further benefit of 3 hops exceeds 10%). In two instances with the highest growth rates, 4-hop provides further benefits, 12.8, 14.7%, and 2, 3 hops incur greater costs compared to the previous hop, while 4-hop attains lower cost in one instance and greater cost in another instance. Thus, the choice for the right number of hops can be 2, 3, or 4 depending on the affordability of the additional costs incurred through the increments. These parameter combinations differs from most settings in that 4-hop provides non-negligible further benefits even for the intermediate growth rate range.

In the high growth rate region, further benefits are mostly substantially lower for 1 and 2 hops and substantially higher for 3 and 4 hops compared to 1.8, 11% false negative rates. The costs above the previous hop are positive. Increasing the number of hops beyond 4-hop provides a further benefit lower than 10.2%. Thus, the choice of the hop number will depend on the affordability of the additional costs. And, in many instances in intermediate and high growth rate regions, the principle of diminishing return for further benefits with increase in the number of hops do not hold. Thus, overall, opting for higher number of hops is more beneficial in this extreme scenario.

### Test Result Turnaround Time

In most of the low growth rate region (except the 2 highest growth rates), the minimum further benefit of 1-hop is 75%, while the maximum further benefit of 2-hop is 15%, with median 8.2%. Thus, in this region there is a large separation between benefits attained by 1 and 2 hops, though the further benefit of 2-hop exceeds 10% in some instances. In the 2 instances corresponding to the highest growth rates in the low growth rate region, 2-hop becomes the clear choice, since it provides considerable additional benefit compared to 1-hop (50.3, 74.1%) and incurs considerably lower costs. Increasing the number of hops further attains negligible additional benefit. Thus, the choice can be 1 or 2 hops in the low growth rate region, with greater number of instances that may opt for 2 hops than when turnaround time was 1 day (the default scenario). Again, this phenomenon is consistent with our findings in the earlier subsection of the Results section, that for the same setting, phase B starts at the higher end points of the low growth rate region.

For the intermediate growth rate range, either 2 or 3 hops constitute a clear choice; in this, this resembles the default scenario. But 2-hop constitutes the clear choice in only 3 instances which are the lowest of the growth rates. For these, 2 hop provide considerable additional benefits (55 - 95.3%, median 81.8%) and attains considerably lower costs than 1 hop, and 3 hop provides negligible additional benefit (≤ 0.8%). For the rest of the instances, both 2 and 3 hops provide considerable additional benefits, but 3 hops incurs comparable or lower costs as compared to 2 hops. Thus, 2-hop can be ruled out. The further benefit of 4-hop is at most 7%, thus 4-hop may be ruled out by our criteria. Besides, 4-hop incurs greater costs than 3-hop. Thus, by and large, 3 hops is a clear choice in these.

In the high growth rate region, the further benefits for 5-hop is at most 10.7%, thus, 5-hop can mostly be ruled out by our criteria. The costs above the previous hop are mostly positive. Thus, 2, 3 and 4 hops may be chosen depending on the further benefits and affordability of the additional costs as for the default scenario. Note that further benefits are lower for 1-hop, mostly lower for 2-hop, comparable or lower for 3-hop, higher for 4-hop and 5-hop compared to 1 day turnaround time. For 3 days turnaround time, the principle of diminishing returns for further benefits with increase in the number of hops holds except for a few instances in intermediate and high growth rate regions.

### Starting Point of Contact Tracing

For low growth rates, the trends for when the starting point is at 0.1%, 2-hop contact tracing attains a further benefit lower than 10%. The trends for when the starting point is at 1.1%, 2-hop contact tracing attains a further benefit lower than 10% in all but 2 highest growth rates in the region, and may therefore be ruled out. Thus, 1-hop constitutes a natural choice in these.

In the intermediate growth rate range, when the initial infection levels are 0.1% and 1.1%, the further benefit of 4-hop is lower than 10%, while those of both 2, 3 hops is considerable. Thus, the choice ought to be between 2, 3 hops depending on the affordability of the additional costs of 3 hops over 2 hops. The high growth rate region exhibits similar trends as the default scenario.

### Level of Cooperation with Contact Tracing and Testing: Scenario 2

For low growth rates, as the cooperativity becomes lower, there are more cases where 2-hop provides greater than 10% further benefits. When cooperativity is 100%, the number is 0. When cooperativity is 75%, this happens for 2 highest growth rates in this range; in these 2 instances 2-hop provides 41.9%, 56.5% further benefits with lower costs than 1-hop. Increasing the number of hops beyond 2-hop provide further benefits in the range 0.6 - 3.9%. These two instances resemble those in intermediate growth rate region. When cooperativity is 50%, the further benefits of 2-hop exceed 10% throughout this region (10.7 - 25.2%, median 18.0%), and the costs are mostly lower than 1-hop. Even further benefits attained by increasing the number of hops beyond 2-hop exceeds 10% in several instances (1.3 - 17.9%, median 3.2%). Thus, as cooperativity decreases, for low growth rates, the choice can become 2 or even 3 hops in more and more instances depending on the affordability of the additional costs (note that 2-hop incur lower costs than 1-hop in most cases).

When the growth rate increases, the sparsity in the network that can be traced limits the efficacy of multi-hop contact tracing. In the intermediate and high growth rate regions, for any finite number of hops, the full benefit provided by each hop substantially decreases with decrease in cooperativity. For example, 3-hop contact tracing reduces the outbreak size by 84.6 - 100.0% (median 99.8%) for 100% cooperativity, 26.1 - 95.4% (median 54.9%) for 75% cooperativity, and 3.0 - 44.9% (median 12.3%) for 50% cooperativity. Further benefits by 4 and 5 hops exceeds 10% frequently for 75%: 1) 1.7 - 19.2% (median 13.3%) for 4-hop 2) 0.5 - 17.2% (median 10.3%) for 5-hop. Incrementing the number of hops also mostly continually increase the cost. Thus, the number of hops need to be chosen between 2, 3, 4, and 5 based on the affordability of the additional costs. For 50% cooperativity, due to further decline in the efficacy of multi-hop contact tracing, further benefits of 1) 2 hops is frequently below 10%, 2) 3 hops is mostly below 10%, 3) 4 and 5 hops are below 10%. Thus, the number of hops need to be chosen based on assessment of further benefits and costs. Finally, there is no behavioral difference between intermediate and high growth rate regions for both 75% and 50% cooperativities.

---

**Table S1.** Values of Disease Parameters.

| Parameter | Notation | Value | Reference & Description |
| --- | --- | --- | --- |
| Transmission probability | $\beta$ | [0.1,0.3] | Assumed various scenarios considering [2], [3] |
| Proportion of infections that are asymptomatic | $p_a$ | 0.4 | [4], [5] |
| Mean latency period | $1/\lambda$ | 2 days | Inferred from [6] |
| Mean duration in asymptomatic stage | $1/r_a$ | 7 days | Inferred from [7], [6] |
| Mean incubation period<br>(period between infection and symptom onset) | $1/\lambda + 1/\alpha$ | 5 days | [8], [9] |
| Mean duration from symptom onset to testing | $1/w$ | 4 days | Inferred from [10] |
| Mean duration of symptom onset to recovery/death | $1/w + 1/r_s$ | 14 days | Inferred from [11], [7] |
| Fraction of symptomatics who die | $p_d$ | 0.0065 | [4] |

**Table S2.**
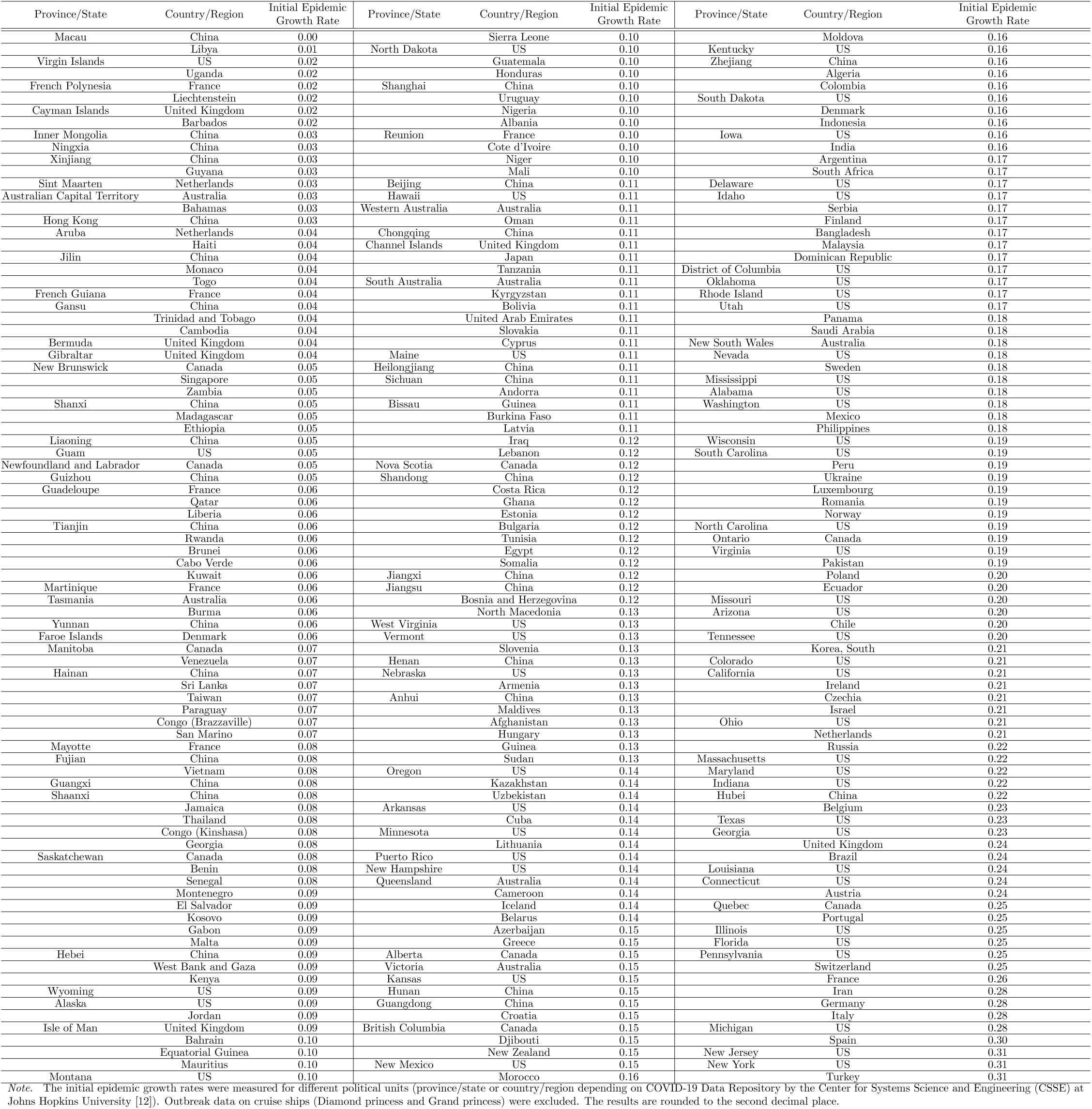
Real-world Initial Epidemic Growth Rates.

**Table S3.** Further Benefit/Cost Across Different Number of Hops (Data-driven Network)

| Intermediate |  |  |  |  |
| --- | --- | --- | --- | --- |
| 1-hop | 2-hop |  | 3-hop |  |
| Benefit | Further Benefit | Further Cost | Further Benefit |  |
| 30.8 - 43.7 (42.1) | 47.6 - 57.7 (53.6) | T: -86.4 - 9.5 (-35.7)<br>Q: -86.2 - 9.3 (-36.0) | $\leq 9.4$ | |

| High |  |  |  |  |  |  |  |
| --- | --- | --- | --- | --- | --- | --- | --- |
| 1-hop | 2-hop |  | 3-hop |  | 4-hop |  | 5-hop |
| Benefit | Further Benefit | Further Costs | Further Benefit | Further Costs | Further Benefit | Further Costs | Further Benefit |
| 10.9 - 29.7 (21.3) | 29.3 - 44.5 (36.4) | T: 69.2 - 162.2 (122.4)<br>Q: 73 - 174.1 (134.2) | 22.5 - 38.4 (29.2) | T: -64.2 - 56.6 (0.1)<br>Q: -64.5 - 57.2 (-0.3) | 3.4 - 13.7 (6.0) | T: 30.1 - 116.9 (64.1)<br>Q: 33.3 - 119.9 (67.8) | $\leq 3.9$ |

*Note.* Results are presented as range and median (in parentheses). ‘T’ stands for Test and ‘Q’ stands for Quarantine. The growth rates for data-driven networks are distributed in the second half of the intermediate range and in the high range.

**Table S4.** Further Benefit/Cost Across Different Number of Hops In *Low* Growth Rates Region (Synthetic Networks)

| Settings |  | 1-hop | 2-hop |  | 3-hop |  |
| --- | --- | --- | --- | --- | --- | --- |
|  |  | Benefit | Further Benefit | Further Cost | Further Benefit | Further Cost |
| Default | | 85.0 - 99.9 (97.8) | $\leq 4.3$ | | | |
| Cooperativity<br>(Scenario 1) | 75% | 83.6 - 99.9 (97.3) | $\leq 5.6$ | | | |
| | 50% | 82.0 - 99.8 (97.2) | $\leq 6.7$ | | | |
| False Negative | 1.8% | 84.6 - 99.9 (99.2) | $\leq 4.6$ | | | |
|  | 58% | 65.1 - 97.5 (90.2) | 2.4 - 13.3 (9.5) |  |  |  |
| | | 29.2, 35.8 | 62.4, 70.5 | T: -98.8, -92.9<br>Q: -98.7, -92.8 | $\leq 1.8$ | |
| False Positive | 0.7% | 84.3 - 99.9 (97.7) | $\leq 4.6$ | | | |
| Turnaround<br>Time | 3 days | 75.8 - 98.7 (91.4) | 1.2 - 15 (8.2) |  |  |  |
| | | 25.7, 49.6 | 50.3, 74.1 | T: -99.5, -99.4<br>Q: -99.4, -99.4 | $\leq 0.06$ | |
| Starting Point of<br>Contact Tracing | 0.1% | 14.0 - 99.6 (94.7) | $\leq 8.2$ | | | |
| | 1.1% | 89.0 - 98.2 (97.1) | $\leq 5.4$ | | | |
| | | 69.7, 75.1 | 23.8, 28.5 | T: -84.1, -77.5<br>Q: -82.8, -76.2 | $\leq 0.8$ | |
| Cooperativity<br>(Scenario 2) | 75% | 77.1 - 98.6 (93.8) | 1.2 - 9.8 (5.4) |  |  |  |
| | | 42.4, 52.3 | 41.9, 56.5 | T: -96.6, -78.6<br>Q: -96.5, -78.9 | $\leq 3.9$ | |
|  | 50% | 4.8 - 84.3 (68.1) | 10.7 - 25.2 (18.0) | T: -74.8 - 13.3 (-52.8)<br>Q: -73.8 - 13.7 (-51) | 1.3 - 17.9 (3.2) | T: -14.3 - 32.3 (-4.6)<br>Q: -14.5 - 32.1 (-5.3) |

*Note.* Results are presented as range and median (in parentheses). ‘T’ stands for Test and ‘Q’ stands for Quarantine.

**Table S5.**
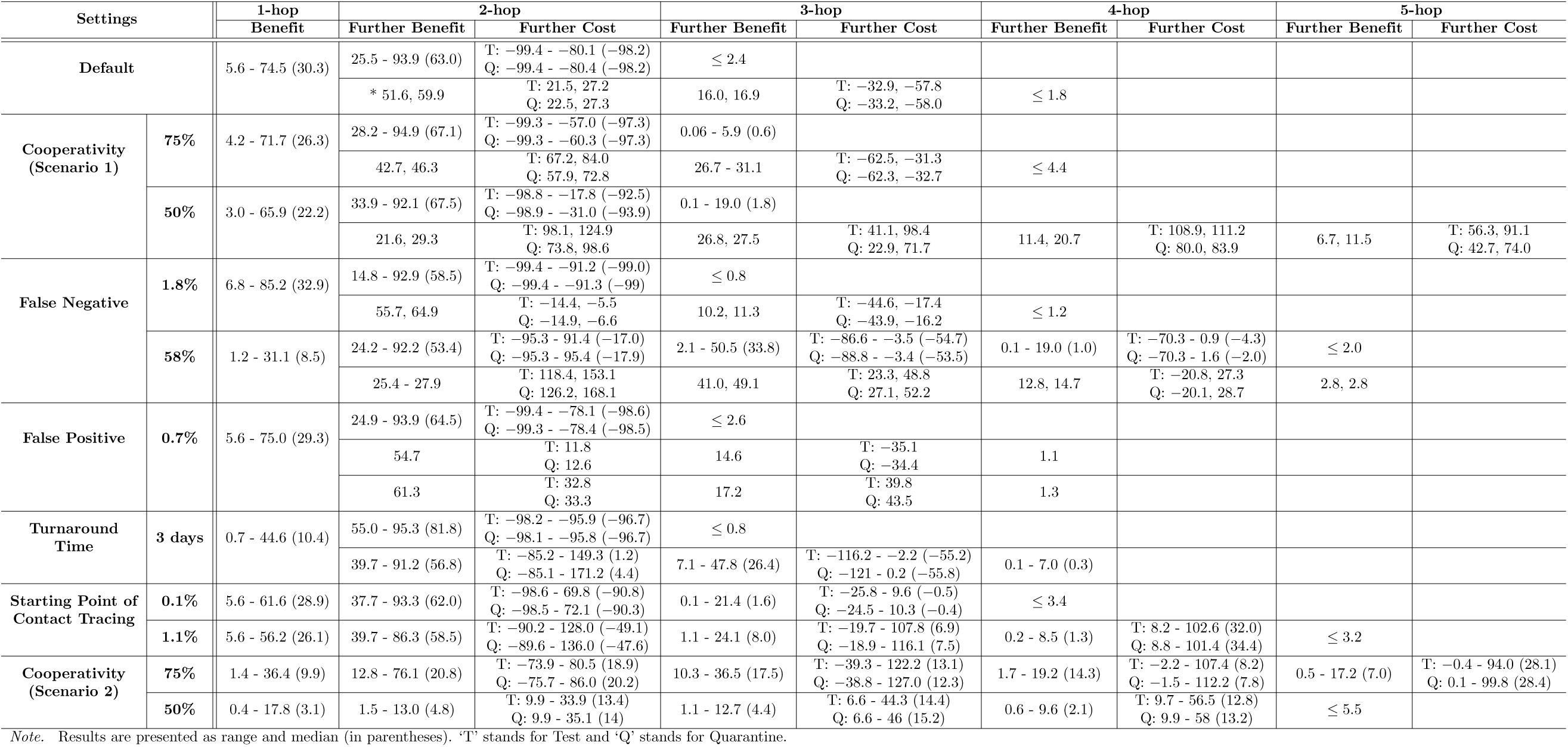
Further Benefit/Cost Across Different Number of Hops In *Intermediate* Growth Rates Region (Synthetic Networks)

**Table S6.**
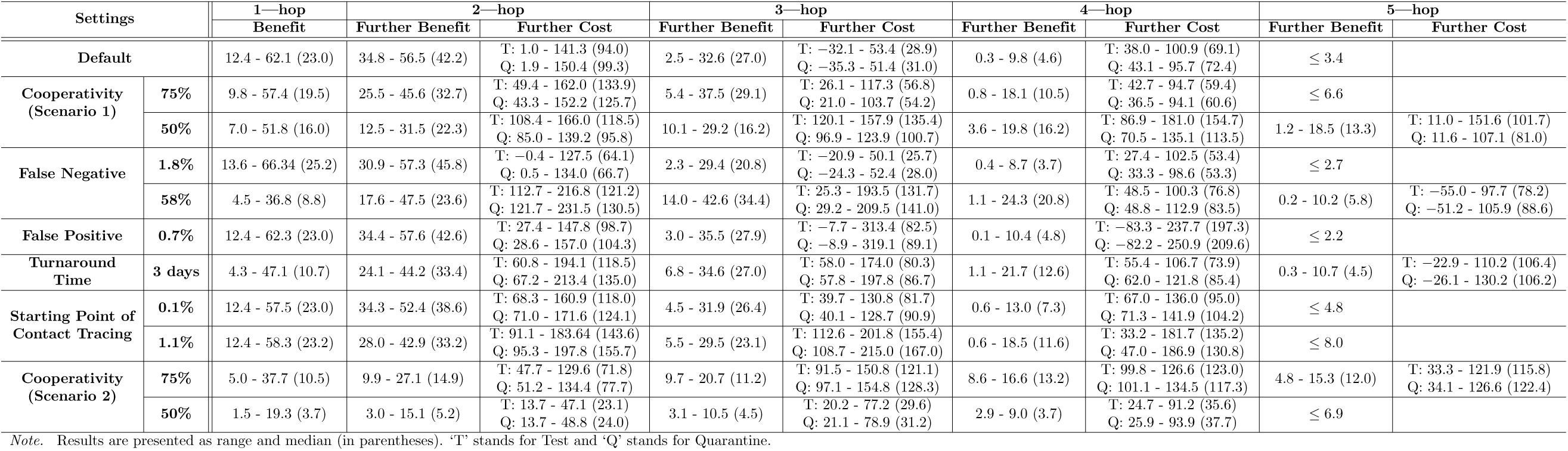
Further Benefit/Cost Across Different Number of Hops in *High* Growth Rates Region (Synthetic Networks)

## References

1. The U S Centers for Disease Control and Prevention (CDC). Contact Tracing for COVID-19; 2020. https://www.cdc.gov/coronavirus/2019-ncov/php/contact-tracing/contact-tracing-plan/contact-tracing.html (accessed 1 December 2020).

2. He X, Lau EH, Wu P, Deng X, Wang J, Hao X, et al. Temporal dynamics in viral shedding and transmissibility of COVID-19. Nature medicine. 2020;26(5):672–675.

3. Ferretti L, Wymant C, Kendall M, Zhao L, Nurtay A, Abeler-Dörner L, et al. Quantifying SARS-CoV-2 transmission suggests epidemic control with digital contact tracing. Science. 2020;368(6491).

4. Bouffanais R, Lim S. Cities-try to predict superspreading hotspots for COVID-19. Nature. 2020;583(7816):352–355.

5. Chungnam Center for Infectious Diseases Control and Prevention, Korea Centers for Disease Control and Prevention (KCDC). Investigation of COVID-19 outbreaks through Zumba dance classes in Korea. Public Health Weekly Report. 2020;13(13):726–736.

6. Lewis D. Why many countries failed at COVID contact-tracing-but some got it right. Nature. 2020;588(7838):384–387.

7. Thai PQ, Rabaa MA, Luong DH, Tan DQ, Quang TD, Quach HL, et al. The first 100 days of severe acute respiratory syndrome coronavirus 2 (SARS-CoV-2) control in Vietnam. Clinical infectious diseases. 2021;72(9):e334–e342.

8. Van Tan L. COVID-19 control in Vietnam. Nat Immunol. 2021; p. 261–261.

9. Baharudin H. More than 1,100 users have deregistered from TraceTogether: Vivian. The Straits Times. 2021;.

10. Watts DJ, Strogatz SH. Collective dynamics of ‘small-world’networks. Nature. 1998;393(6684):440–442.

11. Madewell ZJ, Yang Y, Longini IM, Halloran ME, Dean NE. Household Transmission of SARS-CoV-2: A Systematic Review and Meta-analysis. JAMA network open. 2020;3(12):e2031756–e2031756.

12. Bi Q, Wu Y, Mei S, Ye C, Zou X, Zhang Z, et al. Epidemiology and transmission of COVID-19 in 391 cases and 1286 of their close contacts in Shenzhen, China: a retrospective cohort study. The Lancet Infectious Diseases. 2020;20(8):911–919.

13. Salathé M, Kazandjieva M, Lee JW, Levis P, Feldman MW, Jones JH. A high-resolution human contact network for infectious disease transmission. Proceedings of the National Academy of Sciences. 2010;107(51):22020–22025.

14. Eubank S, Guclu H, Kumar VA, Marathe MV, Srinivasan A, Toroczkai Z, et al. Modelling disease outbreaks in realistic urban social networks. Nature. 2004;429(6988):180–184.

15. Banerjee A, Chandrasekhar AG, Duflo E, Jackson MO. The diffusion of microfinance. Science. 2013;341(6144).

16. Merow C, Urban MC. Seasonality and uncertainty in global COVID-19 growth rates. Proceedings of the National Academy of Sciences. 2020;117(44):27456–27464.

17. Dong E, Du H, Gardner L. An interactive web-based dashboard to track COVID-19 in real time. The Lancet infectious diseases. 2020;20(5):533–534.

18. Arevalo-Rodriguez I, Buitrago-Garcia D, Simancas-Racines D, Zambrano-Achig P, Del Campo R, Ciapponi A, et al. False-negative results of initial RT-PCR assays for COVID-19: a systematic review. PloS one. 2020;15(12):e0242958.

19. Sung H, Yoo CK, Han MG, Lee SW, Lee H, Chun S, et al. Preparedness and rapid implementation of external quality assessment helped quickly increase COVID-19 testing capacity in the Republic of Korea. Clinical chemistry. 2020;66(7):979–981.

20. Sung H, Han MG, Yoo CK, Lee SW, Chung YS, Park JS, et al. Nationwide external quality assessment of SARS-CoV-2 molecular testing, South Korea. Emerging infectious diseases. 2020;26(10):2353.

21. Görzer I, Buchta C, Chiba P, Benka B, Camp J, Holzmann H, et al. First results of a national external quality assessment scheme for the detection of SARS-CoV-2 genome sequences. Journal of Clinical Virology. 2020;129:104537.

22. Matheeussen V, Corman VM, Mantke OD, McCulloch E, Lammens C, Goossens H, et al. International external quality assessment for SARS-CoV-2 molecular detection and survey on clinical laboratory preparedness during the COVID-19 pandemic, April/May 2020. Eurosurveillance. 2020;25(27):2001223.

23. The U S Centers for Disease Control and Prevention (CDC). Overview of Testing for SARS-CoV-2 (COVID-19); 2021. https://www.cdc.gov/coronavirus/2019-ncov/hcp/testing-overview.html (accessed 30 March 2021).

24. Brockmeier EK. Can contact tracing stop the spread of COVID-19? Penn Today. 2020;.

25. Worldometer; 2021. Available at https://www.worldometers.info/coronavirus/.

26. Firth JA, Hellewell J, Klepac P, Kissler S, Kucharski AJ, Spurgin LG. Using a real-world network to model localized COVID-19 control strategies. Nature Medicine. 2020; p. 1–7.

27. Chen SC. Taiwan’s experience in fighting COVID-19. Nature Immunology. 2021;22(4):393–394.

28. O’Leary N, Scally D. Covid-19: How Asian countries got contact-tracing right while European states are struggling. The Irish Times. 2020;.

29. Koob D. 20% Of COVID-19 Positive Individuals Not Cooperating With Montgomery County Contact Tracers, Officials Say. CBS Philly. 2020;.

30. Riley S, Ferguson NM. Smallpox transmission and control: spatial dynamics in Great Britain. Proceedings of the National Academy of Sciences. 2006;103(33):12637–12642.

31. Ralph P. Pennsylvania’s new coronavirus contact tracing app now available. PhillyVoice. 2020;.

32. Aleta A, Martín-Corral D, y Piontti AP, Ajelli M, Litvinova M, Chinazzi M, et al. Modelling the impact of testing, contact tracing and household quarantine on second waves of COVID-19. Nature Human Behaviour. 2020;4(9):964–971.

33. Worby CJ, Chang HH. Face mask use in the general population and optimal resource allocation during the COVID-19 pandemic. medRxiv. 2020;.

34. Barabási AL, Albert R. Emergence of scaling in random networks. Science. 1999;286(5439):509–512.

35. Newman ME. Mixing patterns in networks. Physical review E. 2003;67(2):026126.

36. Staples PC, Ogburn EL, Onnela JP. Incorporating contact network structure in cluster randomized trials. Scientific reports. 2015;5(1):1–12.

## References

1. Ralph P. Pennsylvania’s new coronavirus contact tracing app now available. PhillyVoice. 2020;.

2. Madewell ZJ, Yang Y, Longini IM, Halloran ME, Dean NE. Household Transmission of SARS-CoV-2: A Systematic Review and Meta-analysis. JAMA network open. 2020;3(12):e2031756–e2031756.

3. Bi Q, Wu Y, Mei S, Ye C, Zou X, Zhang Z, et al. Epidemiology and transmission of COVID-19 in 391 cases and 1286 of their close contacts in Shenzhen, China: a retrospective cohort study. The Lancet Infectious Diseases. 2020;20(8):911–919.

4. The U S Centers for Disease Control and Prevention (CDC). COVID-19 Pandemic Planning Scenarios. 2020;.

5. Oran DP, Topol EJ. Prevalence of Asymptomatic SARS-CoV-2 Infection: A Narrative Review. Annals of Internal Medicine. 2020;.

6. Ma S, Zhang J, Zeng M, Yun Q, Guo W, Zheng Y, et al. Epidemiological parameters of coronavirus disease 2019: a pooled analysis of publicly reported individual data of 1155 cases from seven countries. Medrxiv. 2020;.

7. Byrne AW, McEvoy D, Collins AB, Hunt K, Casey M, Barber A, et al. Inferred duration of infectious period of SARS-CoV-2: rapid scoping review and analysis of available evidence for asymptomatic and symptomatic COVID-19 cases. BMJ Open. 2020;10(8). doi:10.1136/bmjopen-2020-039856.

8. Linton NM, Kobayashi T, Yang Y, Hayashi K, Akhmetzhanov AR, Jung Sm, et al. Incubation period and other epidemiological characteristics of 2019 novel coronavirus infections with right truncation: a statistical analysis of publicly available case data. Journal of clinical medicine. 2020;9(2):538.

9. Lauer SA, Grantz KH, Bi Q, Jones FK, Zheng Q, Meredith HR, et al. The incubation period of coronavirus disease 2019 (COVID-19) from publicly reported confirmed cases: estimation and application. Annals of internal medicine. 2020;172(9):577–582.

10. Biggerstaff M, Jhung MA, Reed C, Fry AM, Balluz L, Finelli L. Influenza-like illness, the time to seek healthcare, and influenza antiviral receipt during the 2010–2011 influenza season—United States. The Journal of infectious diseases. 2014;210(4):535–544.

11. The U S Centers for Disease Control and Prevention (CDC). COVID-19 Pandemic Plannin Scenarios. 2020;.

12. Dong E, Du H, Gardner L. An interactive web-based dashboard to track COVID-19 in real time. The Lancet infectious diseases. 2020;20(5):533–534.

